# Complexity of coupled behaviour-disease models and their relative performance against empirical data

**DOI:** 10.64898/2026.07.23.26358796

**Authors:** Sefah Frimpong, Chris T. Bauch

## Abstract

The initial response of populations to the SARS-CoV-2 virus reduced the incidence of COVID-19 cases. However, this success was shorted lived once most populations relaxed most restrictions, resulting in an increase in infections. This feedback contributed to additional pandemic waves. The temporal unfolding of behavioural changes in populations present a challenge to mathematical models for disease dynamics. Coupled behaviour-disease models with varying levels of complexity accounting for several factors have been used to capture behavioural dynamics and SARS-CoV-2 transmission, with varying results. To study the impact of model complexity on the predictive power of models, here we formulate five coupled behaviour-disease models with varying structure and number of parameters. We fit the models to SARS-CoV-2 infection incidence and stringency of control interventions from five European countries in the first wave, and study how well these fitted models predict the second wave. We show that models with more parameters do not necessarily have a greater ability to explain and predict key features of a pandemic wave. Hence, our results show that a relatively simple coupled behaviour-disease model with important parameters can do an adequate job of providing information about the pandemic wave. Additionally, our findings show that complex models can be country-specific, working better for some countries and poorly for others. We conclude that modellers should not always opt for the most complicated possible models, if the data do not support their use.

## Introduction

Mathematical models have become useful tools for understanding the dynamics of infectious disease spread within a population^1,2^. These models have benefited from several contributions since the foundational compartmental model was formulated by Kermack and McKendrick from 1927 to 1933^3–5^. These contributions have resulted in a better understanding of disease dynamics in the context of data availability and improved screening processes^6,7^. Likewise, social dynamics, environmental changes, climate conditions, migration, urbanisation, and many other factors have now been identified as elements that *“manipulate”* the dynamics of disease outbreaks^8,9^. Although these factors have added to the complexity of epidemiological models, they have greatly contributed to new discoveries, better decision-making processes, and the implementation of mitigation strategies, resulting in reduced infection and death rates, as well as the formulation of public health policies^10–13^. Aside from the benefits to epidemiological studies, mathematical models have facilitated the design and implementation of experimental scientific studies involving either living organisms or chemicals, as well as curated data collection^14^ involving human subjects^15^.

One form of adding complexity to models has been through the channel of social dynamics, whose impact on disease dynamics cannot be overlooked^16^. It has become evident that the dynamics of a disease influence social dynamics, which, in turn, also affect the dynamics of the disease – illustrating a feedback mechanism. Such models can be used to study the influence of non-pharmaceutical interventions such as social distancing, mask-wearing, and international and local border controls^17–20^, as well as pharmaceutical interventions such as vaccination and antiviral drug administration^21,22^, factors like fear, opinions, rumours, or information spread^23^, and the effects of social norms^24^. These are characterised by a feedback loop between the disease system and the intervention strategy (which affects social dynamics). The COVID-19 pandemic illustrated a strong feedback loop between social behaviour and the disease^17^. Many mathematical models have attempted to capture this interplay in various forms, largely by considering different approaches to modelling the behavioural responses of the population.

Models of the evolution of social dynamics formulation have been based on at least two main factors^25^ in a broad context: belief-based and prevalence-based, while others include incidence-based (new cases instead of prevalence)^26^, and the payoff(utility) individuals derive from their decisions^27^. Prevalence-based behaviour changes take different forms based on current prevalence, the current and past history of disease prevalence^28^, and media reported prevalence instead of actual^29^. Utility-based behaviour can be estimated as a function of prevalence, as a fixed rate independent of current prevalence, or through social learning(imitation) influenced by the structure of society^27,30,31^. The formulation of these models, coupled with disease models, results in coupled behaviour-disease models that take different structural forms of the interaction between the social system and the disease system. One such structure is interacting networks^32,33^, which consist of at least two layer networks where one network layer represents disease transmission, and the second network or other network layers could represent social communication for the influence of information or behaviour in a scale-free or random nature^34–36^.

A simple yet useful way to formulate a coupled behaviour-disease model is to use the concept of compartmentalisation of the population^37^. Most network models make use of the foundational concept of this approach^33^. In this modelling technique, the complete system is made up of two subsystems: a disease subsystem and a behaviour dynamics subsystem. The disease (epidemiological) subsystem divides the population into groups based on their status: susceptible, exposed, infectious, and recovered, where transitions from one state to another are described by ordinary differential equations^27,38^. The behaviour dynamics subsystem can take two forms: i) a further subdivision of disease groups based on their beliefs, risk perception, or adherence to interventions such as mitigating susceptible and non mitigating susceptible groups^39,40^; ii) a game theoretical model of behaviour response when individuals engage in a game of decision-making^33,41^. The two subsystems are coupled mostly through the transmission (contact) rate parameter (modelled as a function of behavioural factors such as the number of individuals engaging in mitigating strategies to decrease disease prevalence, increase the rate of individual awareness, or encourage the adoption of protective interventions with respect to prevalence level or information spread), as is the case with SARS-CoV-2 transmission^42^. Behavioural models that are modelled using game theoretical frameworks, where individuals are primarily interested in maximising their utility, operate on the notion that individuals make decisions based on several factors and can change their decisions over time as the epidemic unfolds^33^.

One challenging question most researchers face at this stage is how many factors to consider in the model formulation and which ones hold significant importance in arriving at the research objective. More complex models possess the ability to capture intricate patterns in the systems^33^; however, the purpose, availability of data, and contextual structure guide the complexity of models^11,43,44^. While some consider a simple behaviour-dependent transmission rate in the context of precautionary measures such as NPIs, others look at simple utility functions, such as the costs of mitigating and not mitigating; some extend this further to include social norms, while others incorporate the homophily of social groups, among many other factors^45,46^. This mostly involves at least two strategies, where one strategy (mitigating) causes a change in disease prevalence and the other maintains a “status-quo” strategy (non-mitigating behaviour). The number of strategies (behaviour types/players) can exceed 2 to *N* finite numbers of behaviours, with utility functions assigned to each strategy based on perceived or expected payoffs (cost or benefit)^47^. These utility functions can take very simple to complex forms. All these considerations contribute to the complexity of the final model, but in the context of the model’s ability to fit empirical data and perform predictions, how important are these?

This brings us to the focus of this study: how complex should models be for the purpose of fitting and prediction? Very few coupled behaviour-disease models are validated against data due to a lack of data on population behaviour. However, the SARS-CoV-2 virus has led to various data collection efforts on the behavioural responses of individuals in many countries. We take advantage of this data to evaluate the predictive powers of five coupled behaviour-disease models with slightly varying behaviour models and different numbers of model parameters. Here, we investigate the potential of these different models to predict the second wave of case notifications during the COVID-19 pandemic, having fitted them to the first wave. We investigate which parameter considerations are significant for this purpose; that is, how the parameters influence the model prediction by working with data from five European nations (Austria, the Netherlands, Portugal, Denmark, and the United Kingdom).

## Results

### Modelling Framework

We consider five coupled behaviour-disease models for SARS-CoV-2 virus transmission in this study, where the structure of coupling the disease model and the behaviour model is the same for four of the models. The disease model is a seasonal SIR compartmental model with three groups: susceptible (S), infected (I), and recovered (R). The susceptible group consists of individuals who have a high probability of infection; the infected group consists of individuals who are hosts of the virus and can transmit it to others; and the recovered group consists of individuals whose health has been restored to its previous state or who are dead as a result of the infection. The disease model remains the same for all five coupled behaviour-disease models. The differences in these models arise from the structure of the behaviour dynamics models, which are formulated using the concept of evolutionary game theory. We consider two behaviour types: mitigator (*M*) and non-mitigator (*N*) — with respect to NPIs within the population in the presence of SARS-CoV-2 virus transmission. Mitigators constitute a section of the population that supports the use of NPIs for controlling the spread of the virus, while non-mitigators are individuals who do not support NPIs. Each of these behaviour types is assigned an expression for perceived payoff, which forms the behaviour model, similar to the replicator equation. Full details of each model can be found in the Methods section. The disease transmission and baseline behaviour-disease coupling framework adapts the model introduced in Ref.^38^, which we extend here to compare multiple behavioural model structures.

We fitted the models to empirical data from 5 European countries (Austria, the Netherlands, Portugal, Denmark, and the United Kingdom) to estimate the model parameters and initial conditions generating the posterior distributions for each model and country. The daily incidence from the model is fitted to the case notifications reported by each country, and the behaviour is fitted to the Oxford Stringency index. The total length of data considered for the study is identified as having two major waves: the first wave (March - May) and the second wave (September - December). The models are fitted to the first portion of the empirical data, which contains the first wave (see Table 1) using the Approximated Bayesian Computation Sequential Monte Carlo (see Methods for details).

**Table 1.** Length of data points for each country with start, cutoff and end dates. Summary of start, cutoff (data points fitted) and end date extracted for five European countries selected for case studies.

| Country | Start Date | End Date | Cutoff Date(Length) | Data Length |
| --- | --- | --- | --- | --- |
| Austria | 25-02-20 | 28-01-21 | 12-09-20 (200) | 339 |
| Denmark | 27-02-20 | 28-01-21 | 14-10-20 (230) | 337 |
| Netherlands | 27-02-20 | 28-01-21 | 14-09-20 (200) | 337 |
| Portugal | 02-03-20 | 28-01-21 | 18-09-20 (200) | 333 |
| United Kingdom | 31-01-20 | 28-01-21 | 07-10-20 (250) | 364 |

**Table 2.** Model, Number of parameters and initial conditions, and model assumptions. Models with corresponding number of parameters to be estimated, and model assumptions.

| Models | Number of parameters (K) | Assumptions |
| --- | --- | --- |
| Model 1 | 12 | Two behaviour types, cost-induced utility functions, imitation of strategies through sampling |
| Model 2 | 15 | Two behaviour types, cost-induced utility functions, accumulation of socio-economic losses, social norm, imitation of strategies through sampling |
| Model 3 | 14 | Two behaviour types, cost-induced utility functions, accumulation of socio-economic losses, imitation of strategies through sampling |
| Model 4 | 14 | Two behaviour types, cost-induced utility functions, existing social norms, imitation of strategies through sampling |
| Model 5 | 13 | Two behaviour types, cost-induced utility functions, non-mitigators can learn about the disease, proportion of mitigators influence it utility |

**Table 3.** Parameter names, range and their sources. Summary table for parameter and initial conditions ranges. *α* is a proportionality constant and “*pop*” is the population size of the respective countries understudy.

| Parameter | Meaning | Range | Source |
| --- | --- | --- | --- |
| $\alpha$ | proportional economic impact of mitigation | 0 – 2 | (Calibrated) |
| $\beta$ | Transmission rate | 0.1 - 0.9 | $\beta = R_0 \times \gamma^{53}$ |
| $b$ | Seasonal transmission amplitude | 0 - 1 | 54 |
| $\phi$ | phase of seasonal transmission | $-90^\circ - 90^\circ$ | 54 |
| $\varepsilon$ | Intervention effectiveness (Models 1 -4) | 0 - 1 | 55 |
| $\delta_2$ | economic impact of mitigation (Model 2) | $1e-08 - 1e-02$ | (Calibrated) |
| $\delta_3$ | strength of social norm (Model 3) | 0 – 5 | (Calibrated) |
| $\delta_4$ | strength of social norm for NPIs (Model 4) | 0.01 – 5 | (Calibrated) |
| $\varepsilon_5$ | Intervention effectiveness (Model 5) | 0 – 1000 | (Calibrated) |
| $1/\gamma$ | Infectious period | 2 - 10 days | 56,57 |
| $\kappa$ | Social learning rate | 100 - 10,000/day | 16,58 |
| $\eta_0$ | Under reporting factor | 1 - 10 | 51 |
| $\lambda$ | Sensitivity decay | 0 - 0.05 | (Calibrated) |
| $c$ | Cost of maintaining measures | 0 - 0.1 | 59 |
| $I_0$ | Initial infection rate | $1/\text{pop} - \alpha(1/\text{pop})$ | (Calibrated) |
| $x_0$ | Initial population support for NPIs | 0 – 1 | (Calibrated) |
| $Ii_0$ | Initial Incidence rate | $0 - \alpha(1/\text{pop})$ | (Calibrated) |

### First Wave Fit

The five coupled behaviour-disease models fit well to the first wave(March - May) of case notifications for Austria, the Netherlands, and Denmark (Figure 1 **A, B, D**). The models 1, 2, and 3 produce a better fit for Portugal compared to models 4 and 5 (Figure 1 **C**). We observe that models 1 and 2 fit the United Kingdom, whereas the other models fit poorly (Figure 1 **E**) based on a time series analysis.

**Figure 1.**
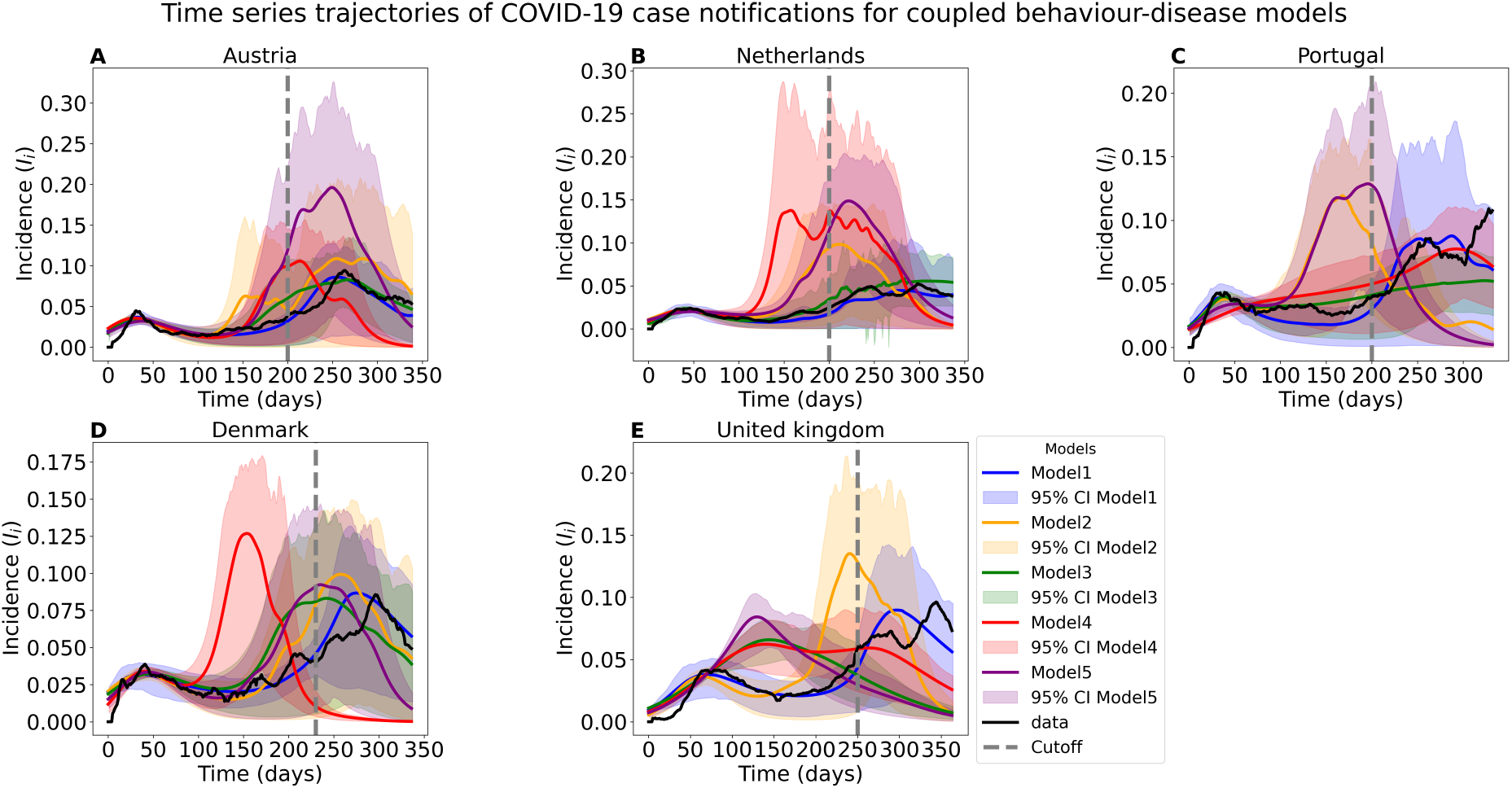
Time series trajectory for model incidence estimation: The five different models fitted output to the first wave of case notifications with the average of 100 parameter set, their 95% credible interval, Cutoff –vertical gray dashed line dividing the training data (first wave) and testing data (second wave) and the performance of their prediction of the empirical data for the second wave (September 2020 - January 2021).

We consider an analysis of the models capability to fit the first wave peak such that the peak magnitude and day are closely matched by our model estimates. We observe from this analysis that the estimated peak magnitude for Austria was lower than the actual for all five models, while the model estimates closely match those for Denmark and the Netherlands. Models 2, 3, and 5 are identified as being below the actual value for Portugal, whereas Model 1 provides a better estimate. We observe in a similar manner that Models 1 and 2 provide good estimates for the United Kingdom compared to Models 3, 4, and 5, which have much higher peak values than the actual(Figure 2 **A**). A corresponding analysis of the peak day using a seven-day interval criterion from the actual peak period reveals that Models 1, 3, and 4 are accurate estimates for Austria. Models 2,3, and 4 are accurate estimates for Denmark; Models 2 and 3 are accurate estimates for the Netherlands, whereas Model 2 is the only accurate estimate for Portugal. Our results reveal that no model accurately estimates the actual peak period for the United Kingdom with respect to our criterion. Nonetheless, when we consider the 95% credible interval, we observe that some of the models meet the criteria for all the countries (Figure 2 **B**).

**Figure 2.**
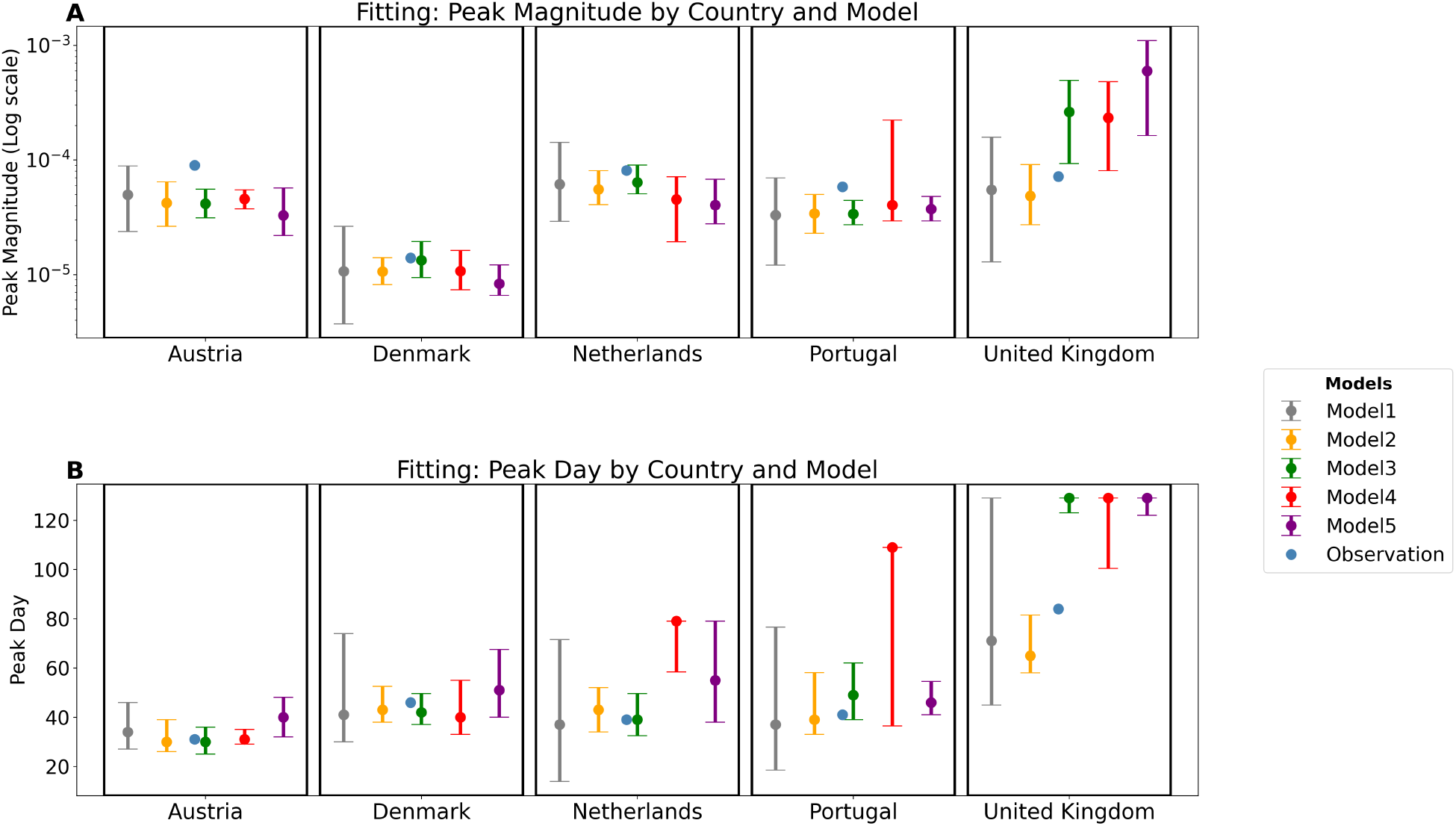
Model estimation of peak magnitude and day after fitting: The peak magnitude and day estimated by each model after fitting to empirical data for the average simulation of 100 parameter set, and their 95% credible interval for each model and country.

Statistically, we employ the Adjusted Akaike Information Criterion (AICc) for evaluating model performance and comparison in two ways: incidence (*I*_*i*_) only and incidence plus behaviour (*I*_*i*_ + *X* ). We observe that Model 1 outperforms all other models for all five countries and has results similar to Model 3 for Denmark and the Netherlands using the incidence only AIC. Model 1 still outperforms all other models for all countries using incidence plus behaviour. In both approaches to calculating the AICc, our results show that Model 1 demonstrates the best goodness-of-fit in comparison to the other models for all five countries (Figure 3). Model 3 yields the second best AIC value on average, based on similar observations. Thus, considering all analyses of estimates from fitting, we can conclude that Model 1 fits the best compared to the other four models.

**Figure 3.**
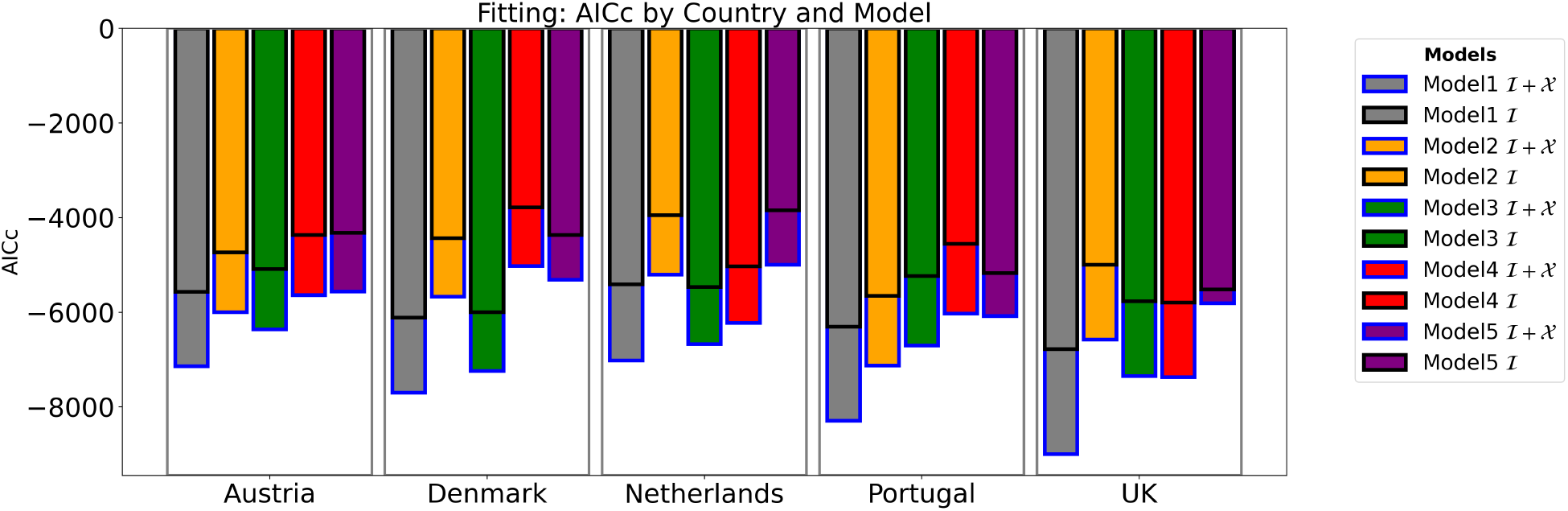
Model calculation of AICc for fitting: AICc calculated for incidence only, and incidence plus behaviour using the average simulation and empirical data for the fitting period for each model and country.

### Second Wave Prediction

We evaluate the predictive capabilities of the coupled behaviour-disease models using the time series trajectories, peak analysis, and statistical metrics. We observe from the time series trajectory that Model 1 predicts the second wave of case notifications in all five countries (Figure 1). Model 3 predicts that, for Austria and the Netherlands (Figure 1 **A**,**B**), Model 2 provides a good prediction for Austria and Denmark (Figure 1 **A**,**D**). All other results show an early and high amplitude second wave prediction. The worst performing model in terms of time series observation is Model 4 for all countries except Portugal (Figure 1 **C**), despite its ability to fit the first wave for those countries (Figure 1 **A**,**B**,**D**,**E**). Although the average simulation of the time series performs poorly, their corresponding 95% credible interval captures the actual case notifications for most of the models.

#### Prediction Peak Magnitude Analysis

The peak analysis provides useful information to assess the performance of the models in terms of prediction. We observe that the 95% credible interval of the predicted peak magnitude for Models 1 to 4 contains the actual peak magnitude for Austria, while Model 5 estimates a high range relative to the actual (Figure 4 **A**). Models 1, 3, and 4 show that their 95% credible interval contains the actual peak magnitude for Denmark, while Models 2 and 5 predict a high range (Figure 4 **A**). For the Netherlands, we identify that Model 3’s prediction range was lower than the actual value, while the other models do contain the actual value. A similar outcome is observed for Portugal, but in this case, Model 4 predicts a much lower range relative to the actual peak magnitude. Lastly, for the United Kingdom(UK), we identify that Models 1, 2, and 4 predict a 95% credible interval that contains the actual peak magnitude, while Models 3 and 5 predict a lower range. Interestingly, we observe that the average predicted peak magnitude values for Austria, Denmark, and Portugal lie in close proximity to the actual values for Models 1 and 3, although Model 1 appears to have a very good approximation for all five countries (Figure 4 **A**).

**Figure 4.**
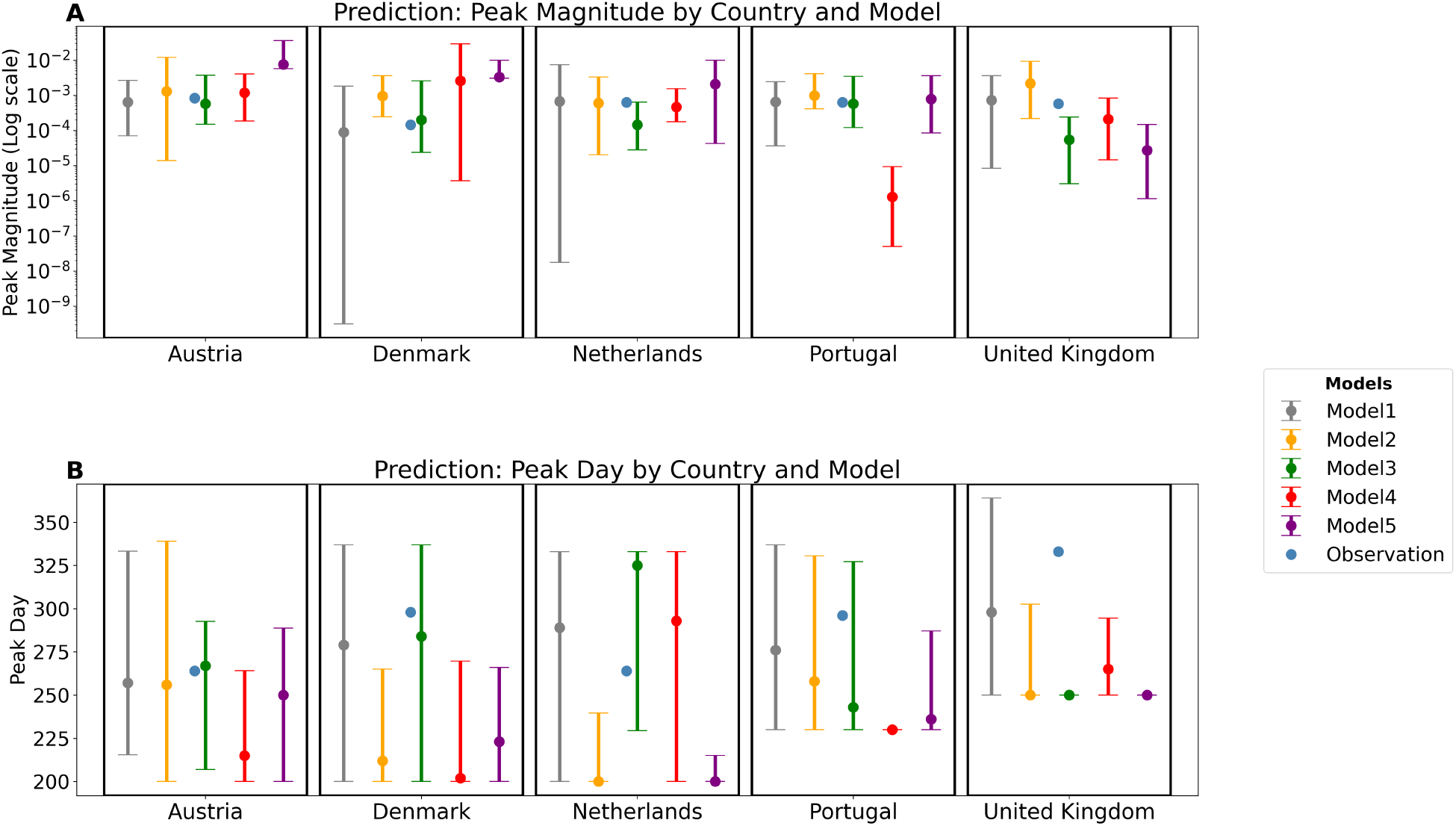
Model estimation of peak magnitude and day for prediction period: The peak magnitude and day estimated by each model for the second wave using the average simulation of 100 parameter set, and their 95% credible interval for each model and country.

#### Prediction Peak Day Analysis

We assess the period when the peak magnitude occurred, first using the 95% credible interval and then a seven-day interval criterion. Our models estimate for the 95% credible interval show that, for Austria’s case notification, Models 1,2,3, and 5 predict a range that contains the actual peak period, while Model 4 predicts a slightly lower range. For Denmark, we observe that Models 1 and 3 provide accurate predictions in terms of their 95% credible interval, while the other models predict much lower intervals compared to the actual values for Denmark. Models 1, 3, and 4 have predicted intervals that align with the actual values for the Netherlands, whereas Models 2 and 5 do not. Models 1 to 3 provide good interval predictions that contain the 95% credible interval for Portugal, while Models 4 and 5 predict lower intervals. We observe that, for the United Kingdom, only Model 1 predicts the 95% credible interval that contains the actual peak period, while the others predict lower intervals (Figure 4 **B**). Now, using a seven-day criterion, we find that the actual peak day for Austria occurs on day 264 of our time series. We identify that only Model 1 meets this criterion with a 257 day prediction. None of the models is able to provide an accurate prediction for Denmark and the United Kingdom. Model 1 predicts 267 days for the Netherlands, whose actual value is on day 264; Model 3 predicts 293 for Portugal, which has its peak period on day 296. Inductively, we can conclude that Model 1 gives the best prediction across all five countries on average compared to the other four models. However, this conclusion is limited without an accompanying statistical evaluation of their performances.

### Statistical Evaluation

We calculate numerical estimates (AICc and Area) for the purpose of model comparison with reference to the empirical data. We focus on the second portion of the data (second wave) for which our models attempt to predict. We again use the Adjusted Akaike Information Criterion(AICc), which penalises the number of parameters in each model and also serves as a measure of goodness-of-fit. We also calculate the area (Area) between the model(s) predictions and the empirical data of case notifications. In a similar manner to the first wave, using AICc for evaluation and comparison, we consider AICc for the incidence (*I*_*i*_) only and AICc for the incidence plus behaviour (*I*_*i*_ + *X* ). Our results show that Model 1 outperforms the other models for both incidence (*I*_*i*_) and incidence plus behaviour (*I*_*i*_ + *X* ) for all five countries, followed by Model 3 for Austria and Denmark. Model 4 outperforms Model 3 for the Netherlands and the United Kingdom regarding AICc incidence, but we observe that Model 3 outperforms Model 4 when considering both incidence and behaviour for Portugal, with a slight difference (Figure 5).

**Figure 5.**
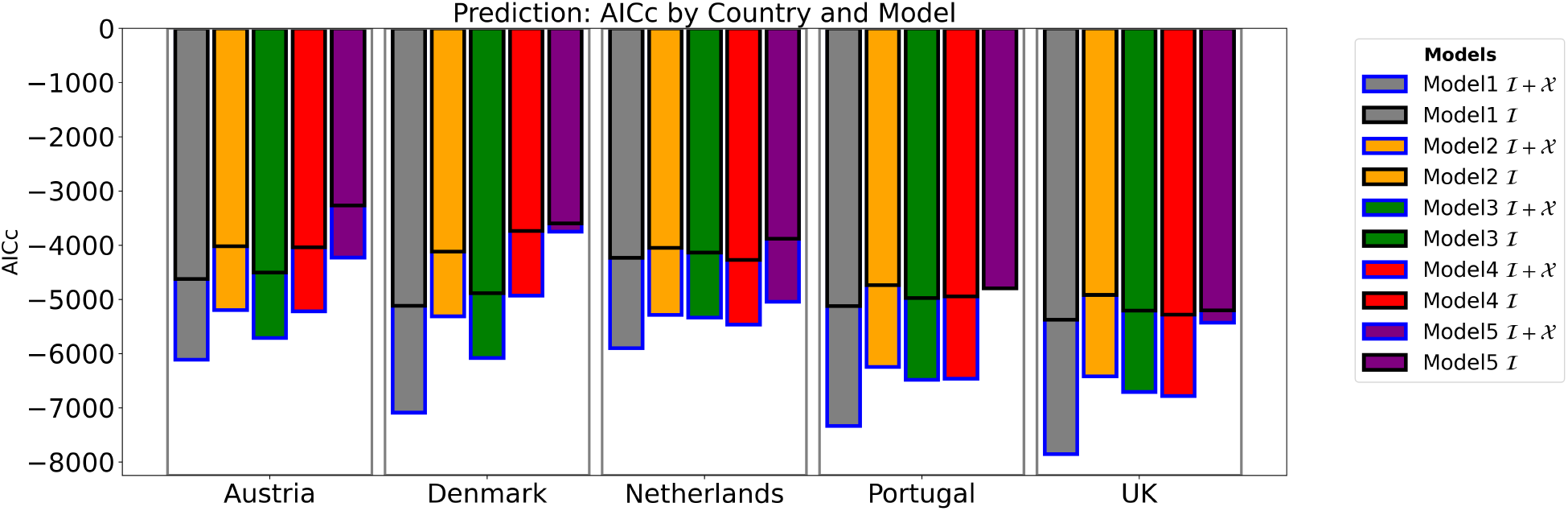
Model calculation of AICc for prediction: AICc calculated for incidence only, and incidence plus behaviour using the average simulation and empirical data for the prediction (September 2020 - January 2021) period for each model and country.

The area is calculated using the residuals between the model output and case notifications. We observe that Model 1 gives a low value for the area for all countries, indicating better performance, except for the Netherlands compared to other models. Model 4 gives a lower value compared to Model 1 for the Netherlands with respect to the area, indicating better performance. Model 3 also predicts the second lowest values of area for Austria, Denmark, and Portugal (Figure 6). Following these results, we can observe that Model 1 predicts the features of the second wave of SARS-CoV-2 the best amongst the five models evaluated in this study.

**Figure 6.**
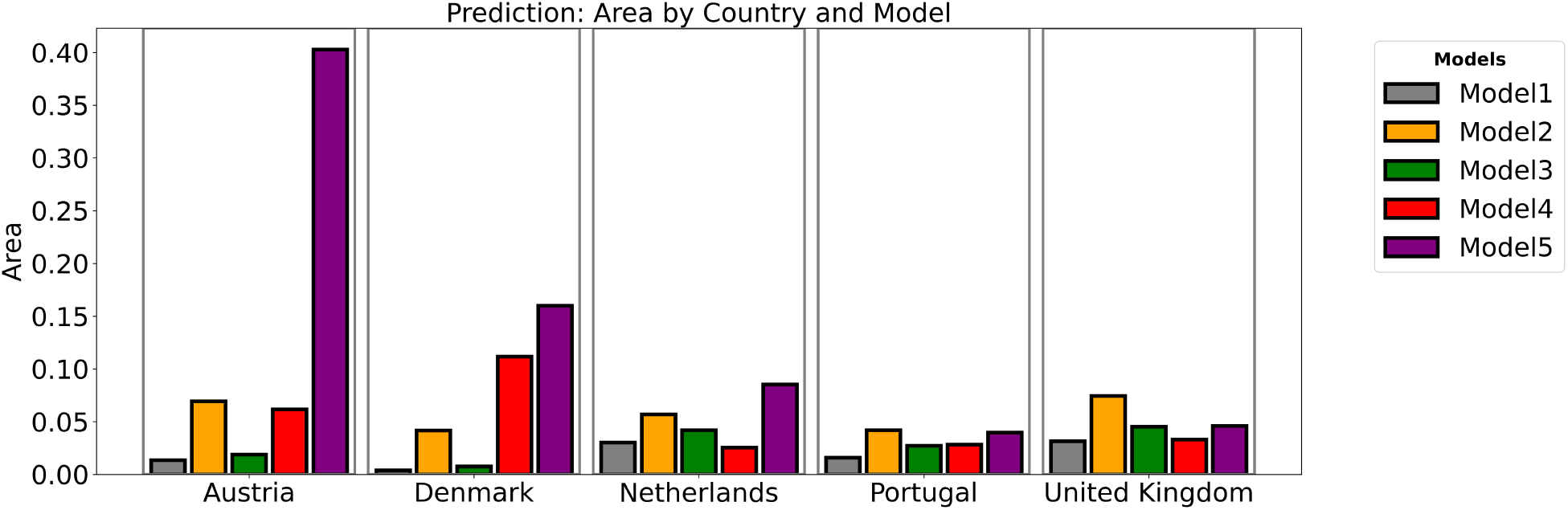
Model calculation of Area for prediction: Area between model prediction of incidences and case notifications calculated using the average simulation and empirical data for the prediction (September 2020 - January 2021) period for each model and country.

### Mitigating Behaviour

The behavioural model is fitted to the Oxford Stringency Index from day 1 to the cutoff (Figure 7, Table 1) for all the models of the five selected European countries. We observe that Model 1 fits well with all the countries, whereas Model 4 fits very poorly. Model 2 also fits poorly for all the countries except Portugal; likewise, Model 3 fits poorly for the Netherlands, Denmark, and the United Kingdom. Model 5 fits quite well in terms of the trajectory of the data for all the countries; however, the estimates are higher than the empirical data. We observe similar results for the prediction of the second portion of the data. Model 1 predicts the behaviour of mitigators in the population with accuracy, while the others perform poorly, except for Model 5, which gives a good prediction for Austria and Portugal compared to the other models. The AICc for prediction affirms our results on the behaviour (Figure 5), indicating that Model 1 is both a good fit and a good predictor of COVID-19 case notifications and changes in population behaviour.

**Figure 7.**
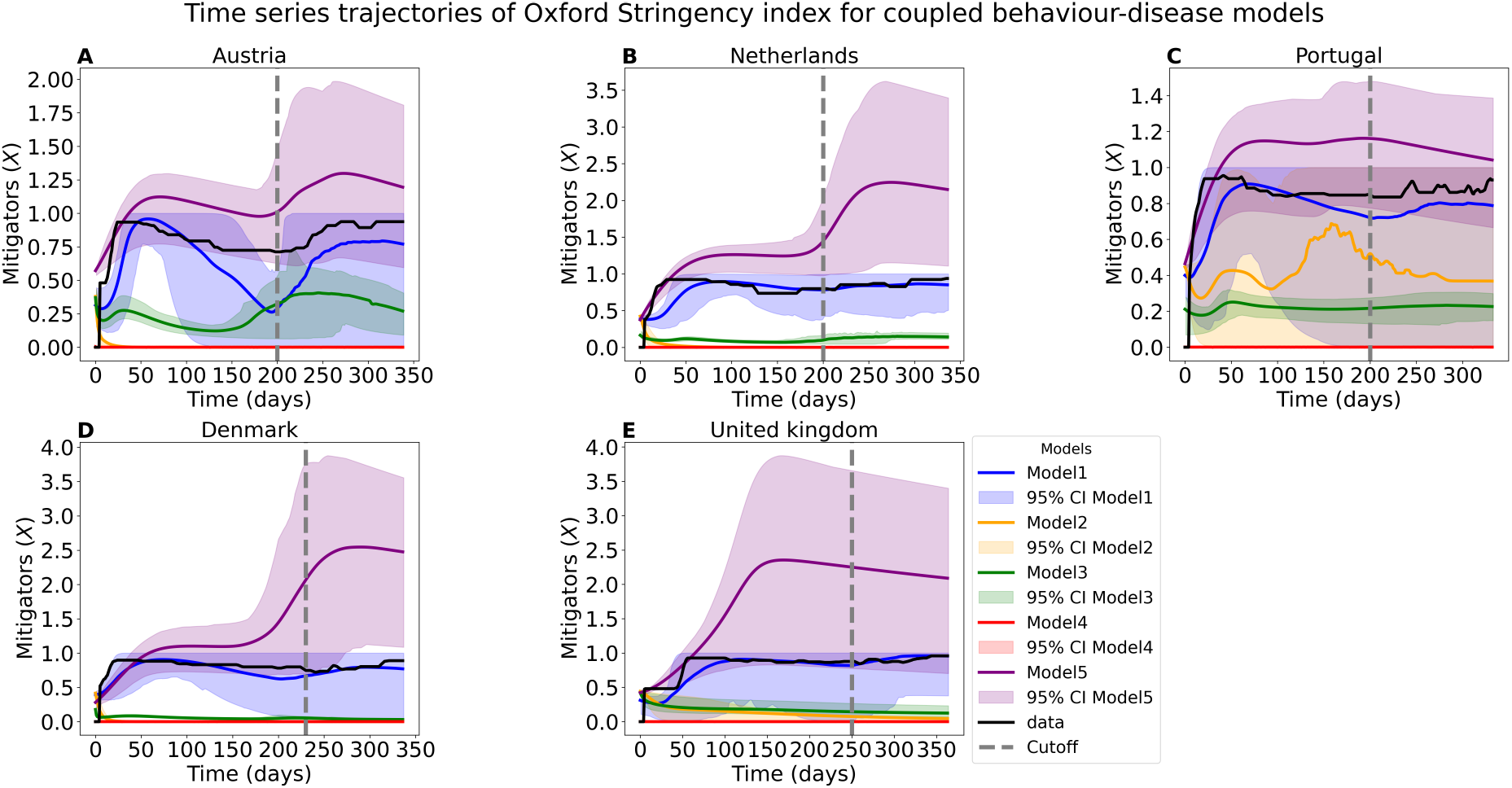
Time series trajectory for model mitigators estimation: The five different models fitted output to the cutoff (gray vertical dash line) of the Oxford Stringency index with the average of 100 parameter set, their 95% credible interval and the performance of their prediction of the second part of the empirical data (September 2020 - January 2021).

## Discussion

In this study, we evaluated the explanatory and predictive abilities of five different coupled behaviour-disease models, from which we established that complex models with more parameters do not necessarily translate to the model’s ability to fit accurately to empirical data and also produce good predictions. Whereas simplified models are able to do so. However, these additional parameters of more complex models do provide useful information about the dynamics of SARS-CoV-2 virus transmission when the models fit well, as our findings reveal in the case of three models (Model 1, 2, and 3). Our findings also reveal that these extra parameters may not be important for prediction purposes; yet, they indicate that certain key parameters are relevant for all the models and demonstrate a significant level of sensitivity.

The usefulness of a coupled behaviour-disease model can be categorised into two major parts: 1) model analysis in the absence of empirical data and 2) model analysis in the presence of empirical data. The former mostly takes a theoretical approach and may involve evaluating model parameters to assess their influence on the model, presenting general ideas that are not verifiable by the study itself since it is conducted in isolation from empirical data. The second part considers models formulated based on the dynamics of the available data. These models can provide very useful insights into the model; however, this is dependent on the accurate estimation of the model parameters. Our results show that, while some parameters are of great importance to the formulation of coupled behaviour-disease models, others make very little contribution to the model’s ability to estimate the empirical data. The simplest of the five models demonstrates that fewer parameters can achieve an accurate fit by obtaining good parameter estimates. These estimates are also easily attainable compared to models with more parameters due to the level of complexity. They also show the possibility of producing good predictions from the simple model, while the more complex models do not produce good predictions for all five countries, but only for a subset. The structure of the model significantly influences its predictability, as our findings reveal when comparing Models 1-4 (similarity in structure) and Model 5, which predicts early and high second waves.

Despite the challenges of the more complex models, we identified that useful information can be extracted from the extra parameters, particularly those that fit well with the empirical model. For instance, this study shows that Models 2 and 3 indicate that socio-economic losses are an important consideration based on empirical data for Austria, the Netherlands, and Denmark. This implies that while mitigation strategies lead to socio-economic losses; as such, the economic impact must be accounted for in models for such countries; although excluding these parameters does not defeat the purpose of fitting and prediction, they provide more information. This consequently reveals that while certain additional parameters may be useful for one set of empirical data, they may not be useful for others. The COVID-19 pandemic was a global health crisis; the dynamics within each country differed, which implies that one model structure may not be a good fit for all. A case-by-case model can prove to be more useful; complex models with more parameters can be case specific and useful in producing desirable results. We clearly see this play out with the empirical data for Portugal and the United Kingdom, where only Model 1 fits and predicts the case notifications accurately. Additionally, this model supports mathematical models with “on-off switches” for certain parameters to suit different scenarios based on the dynamics of empirical data^48^.

This study contributes to the field of coupled behaviour-disease models; nevertheless, there are a number of limitations that could be addressed for further improvement. The first challenge we identify for this study is the fitting algorithm used for all the complex models. Although Approximated Bayesian Computation Sequential Monte Carlo (ABC SMC) is a good parameter estimation algorithm, it may not be suitable for different complex models. Additionally, we assumed the same parameter range for the prior distribution to estimate the posterior distribution for all the models and countries. The different models may need to be sampled from different ranges. Another limitation is the number of fitting sequences that need to be carried out. We considered 30 sequences (iterations), but due to the complexity of the models, more than 30 sequences of fitting may be needed to obtain a good fit and prediction. As we have already established, coupled behaviour-disease models in their simplest form are useful for fitting and prediction compared to the disease model. As such, for future work, we will consider all 13 European countries from our previous studies^38^, perform stratification of the models with the empirical data to identify the best model-data outcome. In doing so, we will explore fitting different models across different ranges, simulating more sequences using the ABC SMC algorithm and other fitting algorithms, such as Gradient Descent.

A further potential limitation of this study is the challenge of parameter identifiability of the models for various countries. We considered the Fisher Information Matrix (FIM), the Bayesian posterior analysis, and sensitivity analysis, but we could not concretely draw a conclusion on this problem. As a future study, an extensive investigation of the practical identifiability of the models for various countries can be undertaken by employing techniques like profile likelihood. This approach holds one parameter at a specific value while it optimises the other model parameters.

In summary, we have illustrated that increasing the complexity of coupled behaviour-disease models by the addition of more parameters does not automatically imply that such models can fit empirical data and produce good predictions. They can, however, provide useful information from the additional parameter(s) included in the models. We also identified that complex models may be data specific and not a one-size-fits-all, revealing the usefulness of some parameters over others. A complex model, in their simplest form, can achieve a similar objective to what a complex model may not in terms of fitting and prediction, although the latter could perform better when the appropriate processes are followed.

## Methods

We consider five coupled behaviour-disease models with a standard SIR model for the disease transmission of the SAR-CoV-2 virus. The disease system is compartmentalised such that we have susceptible, infected, and recovered (dead) groups. The Susceptible group consists of individuals who, when exposed to the virus, can become infected; the infected group is the set of individuals who have come into contact with the virus, have been infected by it, and may be able to transmit the virus to healthy susceptible individuals; and the recovered (dead) group includes individuals who have either recovered from the virus or died as a result of complications from the infection. The disease system remains the same for all five models, but they have different behavioural models and different coupling approaches. We consider that the virus is affected by seasonality as a result of environmental or climatic changes. The seasonality of the virus is viewed as occurring in phases, as the case notifications reported by countries reveal. Thus, the seasonal SIR model equations are formulated as:

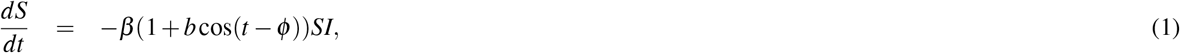

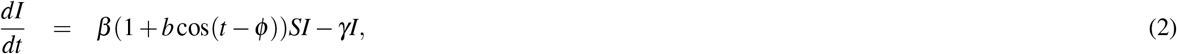

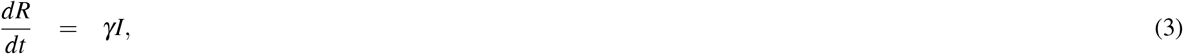

where *S* is the proportion of susceptible individuals (“susceptible”), *I* is the proportion of individuals who are both infected and infectious (“infectious”), and *R* is the proportion of individuals who are no longer infectious (“removed”). *β* is the baseline transmission rate in the absence of non-pharmaceutical interventions, and *γ* is the rate at which an infectious person recovers. *b* represents the amplitude of the seasonal force, and *ϕ* is the phase of seasonality.

## Behaviour Models

The differences and complexity of the coupled behaviour-disease models considered in this study are accounted for by the differences in the behavioural models. Although all five models are built on the concept of evolutionary game theory by considering two behaviour types, their respective utility functions (expressions) differ. There is no doubt that the introduction and implementation of NPIs significantly influence the dynamics of the SARS-CoV-2 virus positively by decreasing infections and SARS-CoV-2 virus related deaths^19^. In spite of these successes, due to the longevity of the survival of the virus, it has become evident that the use of NPIs may not be sufficient to mitigate the spread to low levels to enable the population to return to their pre-COVID-19 state of living. NPIs such as social distancing, isolation, and quarantine, as well as practicing personal hygiene, have become costly for individuals, societies, and nations. This implies that the initial decision to adhere to the use of NPIs had to be reconsidered. At this point, the decision to mitigate or not to mitigate became a game where individuals had to weigh their options and make decisions. The average individual may want to make decisions that are most beneficial to themselves first, then to their household and community, by assessing the costs and benefits of available options (strategies).

We formulate the behavioural model by assuming that only two options (strategies) are available for the individual to choose from. The mitigating (*M*) strategy, where individuals support the use of NPIs, and the non-mitigating (*N*) strategy, where individuals do not support the use of NPIs. We suppose that an individual’s decision on a strategy is not made in isolation from others in the community; that is to say, individuals imitate others in their decision-making since they aim to maximise their utility. We estimate a perceived payoff or utility for each strategy; mitigators payoff (*E*_*M*_) and non-mitigators payoff (*E*_*N*_). If the payoff for mitigators is greater than the payoff for non-mitigators, we expect a change of behaviour from non-mitigators to mitigators and vice versa. We represent the proportion of mitigators by the variable *x*; then the proportion of non-mitigators becomes 1 − *x*. Using the proportional imitation rule (replicator equation), we are able to model the changes in the proportions of both behavioural types, forming our behaviour model, which is then coupled with our seasonal SIR disease model to constitute the coupled behaviour-disease models.

### Base Model

The models 1 to 4 have the same structure with respect to how the disease system is coupled with the behaviour system, although the behaviour has different model structures (forms). Since the behaviour consists of contact precautionary measures, changes in the population influence the transmission potential of the SARS-CoV-2 virus by either enabling or reducing transmission^27,49^.

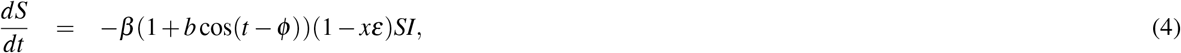

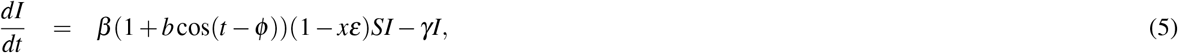

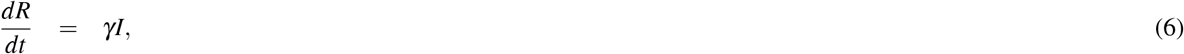

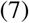

where *x* represents the proportion of mitigators in the population, and *ε* represents the strength of the use of NPIs by mitigators.

We discuss the behaviour model’s structure for models 1 to 4 in the following sections.

### Model 1

This model is the same as in our study of coupled behaviour-disease models, and its ability to produce pandemic waves and demonstrate improved predictability compared to that of a disease model. Full details for such a model can be found in the paper^38^, and the behaviour model equation is:

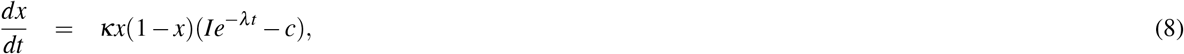

where *κ* is the social learning rate, *λ* is the decay in sensitivity over time as the population learns more about the virus and becomes less afraid of it, and *c* is the cost of maintaining measures. The quadratic term *x*(1 − *x*) represents social learning dynamics, where individuals sample others at some rate and change their opinions based on the utility differences in the brackets.

### Model 2

This model is an extension of model 1, where we include socio-economic losses to influence the cost *c* of mitigation, and the accumulation of socio-economic losses structure changes to account for the strength of the use of NPIs (*ε*). This implies that the perceived payoff for mitigators *E*_*M*_ becomes *E*_*M*_ = −*cL*. Following the same concept of evolutionary game theory with the imitation dynamics of the proportional imitation rule (*E*_*M*_ − *E*_*N*_ = *Ie*^−*λt*^ − *cL*), the model equation for the behaviour dynamics becomes:

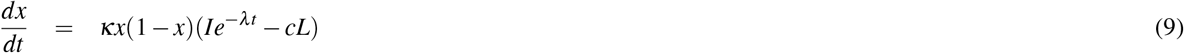

where *L* is a phenomenological representation of accumulated socio-economic losses that obeys

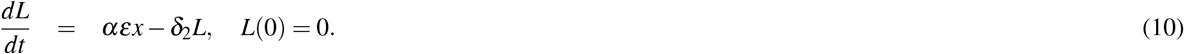

where *α* represents a proportional economic impact of mitigation, and *δ*_2_ represents the strength of the social norm.

### Model 3

This model is a simplification of model 2, where we include socio-economic losses to influence the cost *c*, but the strength of mitigation represented in model 2 is defined as *δ*_2_ = *αε* as a proportional constant of the rate of mitigators. The model equation for the behaviour dynamics becomes:

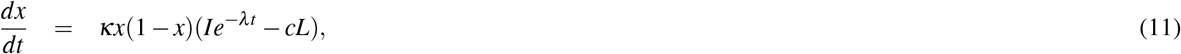

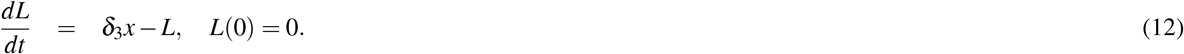

where *δ*_3_ represents the economic impact of the mitigation.

### Model 4

This model introduces social norms in the estimation of the respective perceived payoffs for each behaviour type: mitigators (*E*_*M*_ = *c* + *δ*_4_*x*) and non-mitigators (*E*_*N*_ = −*Ie*^−*λt*^ + *δ*_4_(1 − *x*)); thus (*E*_*M*_ −*E*_*N*_ = −*Ie*^−*λt*^ − *c* + *δ*_4_(2*x*− 1). Following the proportional imitation rule, we obtain the model that accounts for the direct impact of social norms on mitigating strategies.

Socio-economic losses are not included in this model.

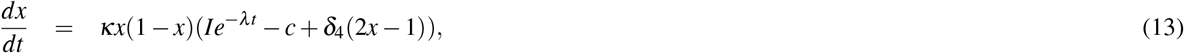

where *δ*_4_(2*x* − 1) represents frequency-dependent social processes and *δ*_4_ represents the strength of existing social norms of NPIs.

### Model 5

This model takes a slightly different structure than the previous four models. The behaviour model still uses evolutionary game theory, but imitation does not follow the proportional imitation rule as in previous models; rather, players base their decisions on the difference in payoffs without necessarily sampling for imitation. The perceived payoff for mitigators is influenced by the proportion of mitigators at each time of decision making, that is, *E*_*M*_ = −*cx*, and the ability of the population to learn about disease transmission also influences the perceived payoff for non-mitigators *E*_*N*_ = *κ* × −*Ie*^−*λt*^. The rate of the proportion of mitigators (*x*) per day is estimated by the difference between the two payoffs (*E*_*M*_ − *E*_*N*_ = *κIe*^−*λt*^ − *cx*). This is then coupled to the disease model by multiplying 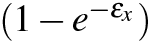 with the transmission rate to account for the increase or decrease in transmissions as a result of behavioural changes. The coupled behaviour-disease model for this is:

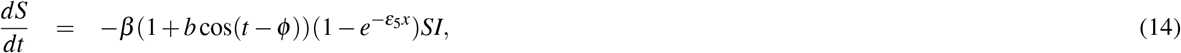

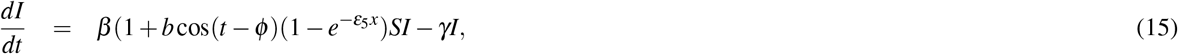

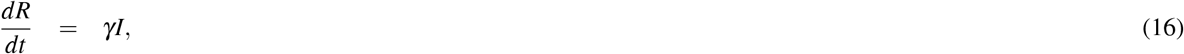

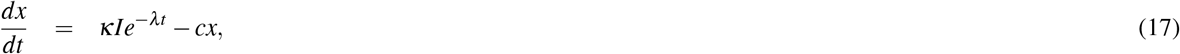

where *ε*_5_ is the strength of the use of NPIs but no longer has to be constrained between 0 and 1 in this model.

### Under-reporting

In a similar fashion to the study Ref. ^38^, we account for under-reporting in all five models, since the chances of under-reporting data are very significant for the COVID-19 pandemic. Social distancing, isolation, and quarantine are effective mitigating strategies for the disease but a disadvantage in terms of data collection, as many asymptomatic and symptomatic cases were not reported for verification. Additionally, the high stress level on hospital facilities and their personnel made it challenging to test all suspected cases for confirmation^50^. To account for this case of under-reporting, we introduce a parameter *η*, which is estimated for this purpose; the reciprocal is taken as the actual value, which is then multiplied by the estimated daily incidences^16^.

### Incidence and Parameterisation

The daily incidence is estimated by formulating the cumulative cases (*C*) for each day (*t*). The daily incidence is evaluated by taking the difference between successive cumulative cases and multiplying it by the under-reporting factor. Since we have two main model structures for coupling the disease system to the behaviour system, we have two expressions for the estimation of the cumulative cases. The first, which we represent as *C*_1−4_, shows that for models 1 to 4, and *C*_5_ represents that for model 5.

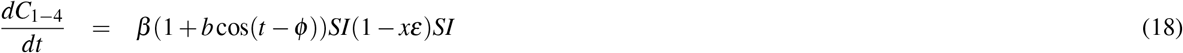

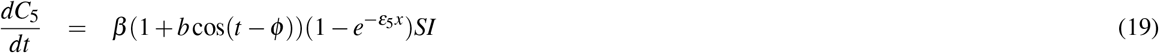

Then the daily incidence for each day (*t*) is estimated as;

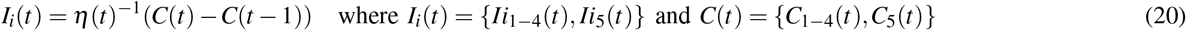

This final expression is then fitted to the daily case notifications reported from each country under study for COVID-19 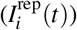 to estimate the model parameters for analysis and prediction^51,52^.

We employ the same parameter estimation approach from Ref. ^38^ of Approximated Bayesian Computation Sequential Monte Carlo (ABC SMC) for each model for the five selected European countries, chosen based on clear first and second waves.

We identify the number of parameters and initial conditions for each model that need to be estimated, as well as those needed for statistical analysis in model evaluation and performance comparison.

To estimate the parameter values for each model and for each country, we assume a range for each parameter, from which we create a uniform prior distribution for sample drawings.

### Parameter Identifiability and Uncertainty

The identifiability of a model is simply the uniqueness of model inputs producing unique outputs. It takes two forms: structural and practical. Structural identifiability can be considered to hold when the model parameters cannot be replaced by fewer parameters^**?**^. Here, Model 2 is structurally unidentifiable since the parameters *α, ε* can be simplified into one parameter–as we have in Model 3–through parameter substitution. Since parameters are estimated from empirical data, we place more emphasis on practical identifiability for this study. Several methods, such as profile likelihood, the Fisher Information Matrix (FIM), Bayesian posterior analysis, and sensitivity analysis, can be employed to detect practical identifiability^60^. We consider the last three for the analysis of our models.

The Fisher Information Matrix measures the amount of information about a set of parameters that can be extracted from a probability distribution given observable data.

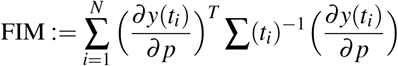

where ∑(*t*_*i*_)^−1^ is the covariance matrix of the measurement noise. The covariance matrix is simply the inverse of the FIM, that is, 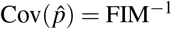. The Bayesian posterior analysis assesses the distribution of the posterior to make judgments on practical identifiability, while the sensitivity also provides additional support for the change in output with respect to changes in model inputs. A significant difference in parameter values corresponding to low changes in output can be classified as having weak identifiability or non-identifiability. Additionally, we consider the correlation matrix for the posterior distribution to assess the relationship between the model parameters. Details of these analyses are presented in the supplementary appendix.

## Data Availability

All data produced in the present study are available upon reasonable request to the authors.

## Authors Contribution

SF conceived the model, and SF established the results. SF draughted the manuscript, and CTB reviewed and edited it. Both authors read and approved the final version of the manuscript. SF accessed and verified the data used in the study and had final responsibility for the decision to submit for publication.

## Acknowledgement

This work was supported by a Discovery Grant to CTB from the Natural Sciences and Engineering Council of Canada (NSERC) [grant number RGPIN-2025-04274]. The funder of the study had no role in study design, data collection, data analysis, data interpretation, writing of the paper, or the decision to submit.

## 1 Supplementary Appendix

### 1.1 Sensitivity Analysis

We evaluate the sensitivity of the parameters in each model for all five countries by considering one parameter set as the model baseline values, which fits fairly well with the empirical data but is not necessarily a good predictor (Figure 8). Using OAT (one-at-a-time) sensitivity analysis and 20% variation of parameter values, we consider three metrics for evaluation: predicted peak day, peak magnitude, and the area between the model output and the case notifications for the various countries. Our results show that all 12 parameters of Model 1 have some level of sensitivity for all three metrics across all five countries. However, we identified that the sensitivity of the area for the Netherlands was very minimal compared to the others, but the peak magnitude indicates that this country was extremely sensitive to all the parameters compared to the other countries. We observe a different case for Model 2, where only five parameters were identified as sensitive for the area across all the countries, with Portugal showing sensitivity only to low variation. With respect to the peak day, we identified 4 parameters for all the countries except Portugal, which identified only one parameter as sensitive. Using the peak magnitude, our results show that Portugal has sensitivity to all parameters except for two, while the remaining four countries identified only 6 parameters as influencing the peak magnitude.

**Figure 8.**
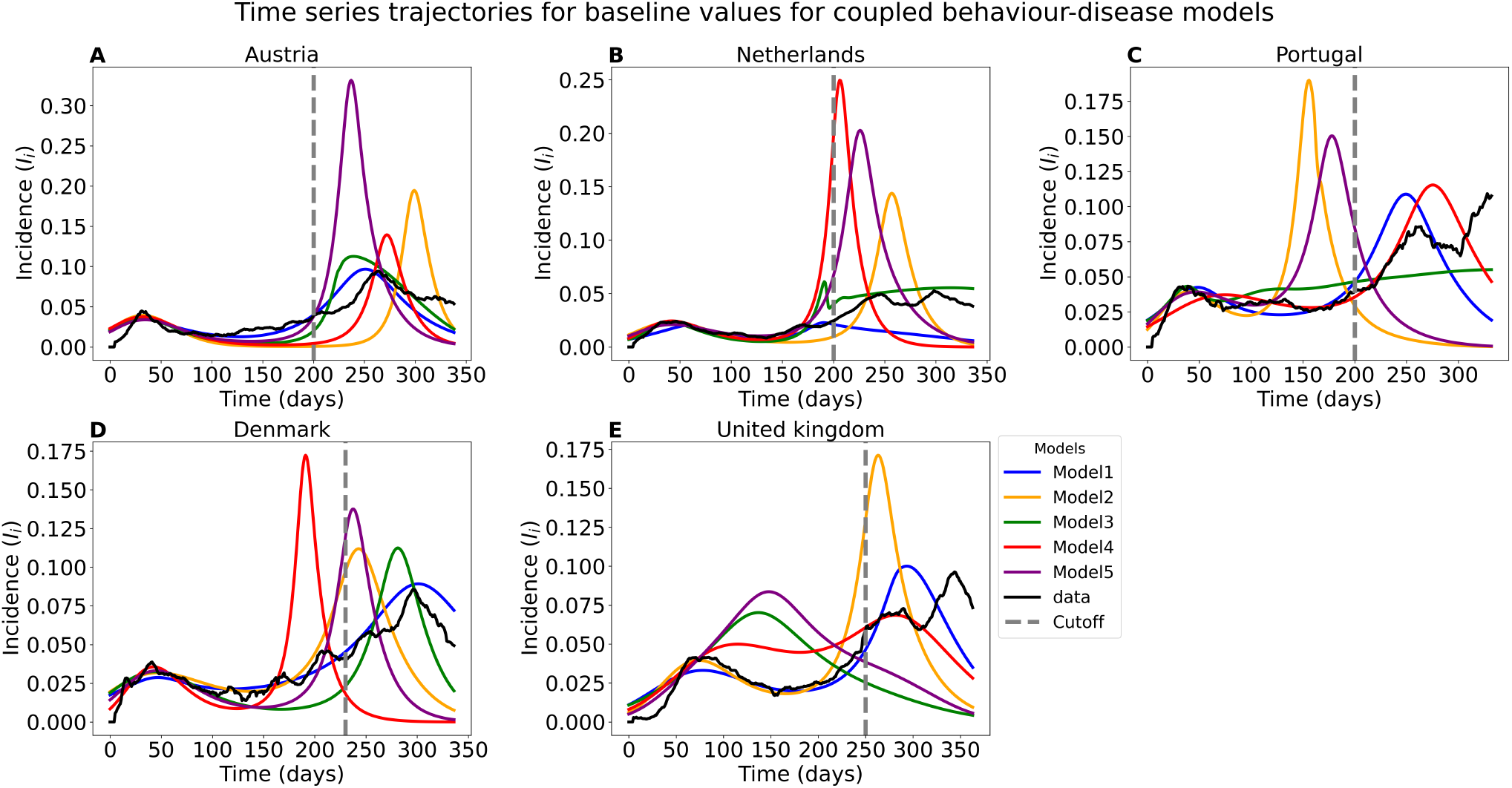
Time series trajectory using selected parameter set as baseline values: A parameter set that fits fairly well to the case notifications is considered as baseline values. They do not have to necessarily have to produce a good prediction.

In a similar approach to previous analyses, Model 3 shows that at least four out of the 14 parameters had no influence on the model with respect to the area for Austria, the Netherlands, and Portugal; Denmark recorded sensitivity for only 6 parameters, while the United Kingdom showed very minimal sensitivity for 3 parameters. The United Kingdom identifies only 2 parameters influencing the peak day, Denmark shows 4 parameters influencing the predicted peak day, Portugal identifies 7 parameters, the Netherlands identifies 9 parameters, while Austria shows that 6 parameters were considered sensitive to the peak day. For the peak magnitude, we observed that 9 parameters were sensitive for Austria, the Netherlands, and Portugal; Denmark was sensitive to 6 parameters, while the United Kingdom was sensitive to 7 parameters.

Model 4 shows a matching number of sensitivities for most countries across the three metrics. We identified 6 parameters that were sensitive for four countries, while Denmark recorded sensitivity for only 3 parameters in the area sensitivity analysis. A similar case is observed for the peak day sensitivity analysis, where Denmark identified only 3 parameters, while the other countries identified 4 parameters. For the peak magnitude sensitivity analysis, we recorded that all the countries identified 6 parameters influencing the magnitude of the peak. We identified some level of parameter sensitivity for Model 5, which showed a degree of uniform results across all five countries. We identified 5 parameters influencing the area sensitivity analysis for all the countries; for peak day, we identified 3 parameters for Portugal and 4 parameters for the other countries. Then, 6 parameters were identified to influence the peak magnitude for all the countries (see Figures 9 -17).

**Figure 9.**
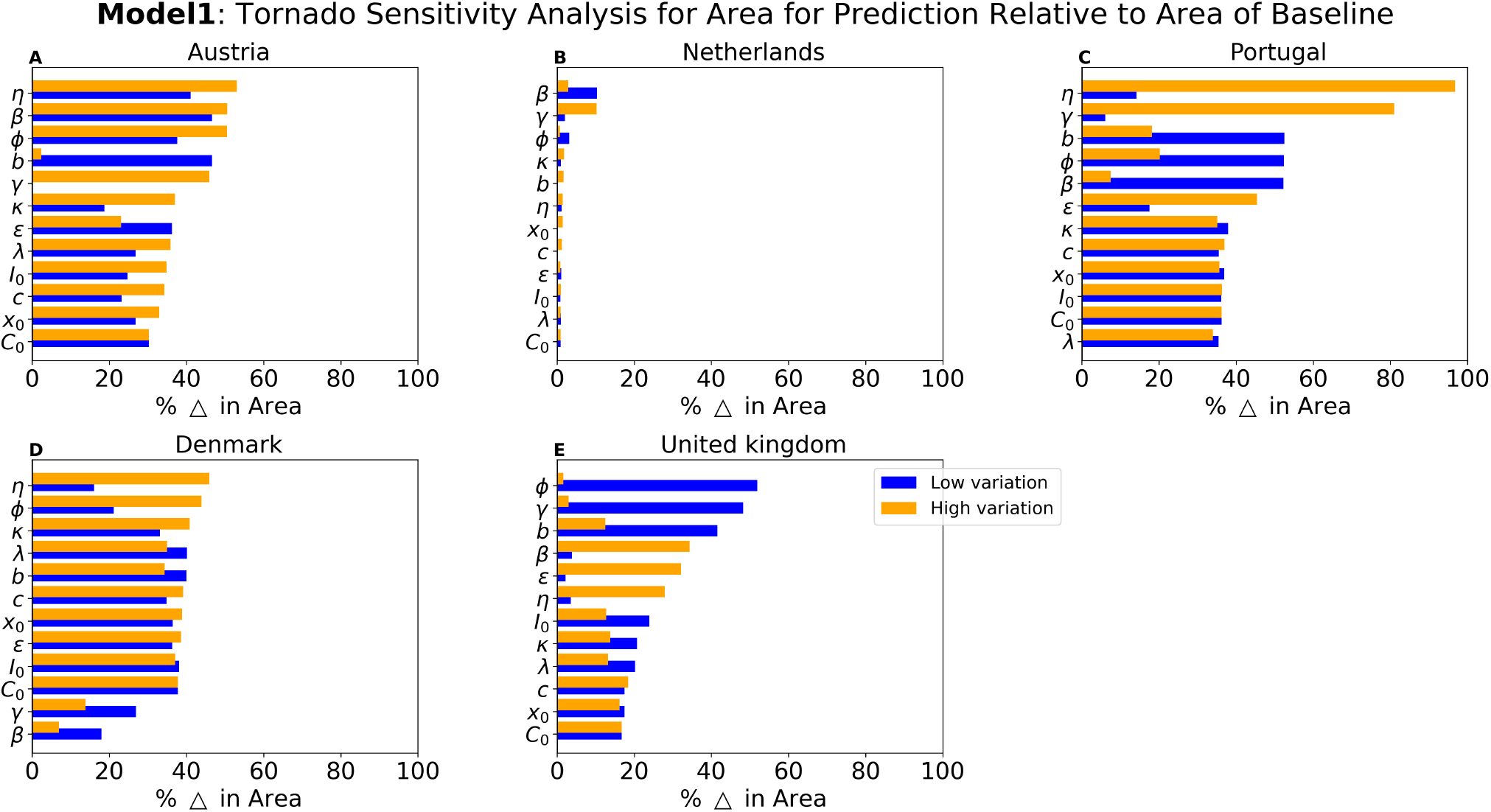
OAT sensitivity analysis using the percentage change in area for Model 1: One-at-a-time sensitivity analysis of each parameter in Model 1 where the area between the model’s incidence prediction and case notifications is calculate. The percentage change is calculated for each variation.

**Figure 10.**
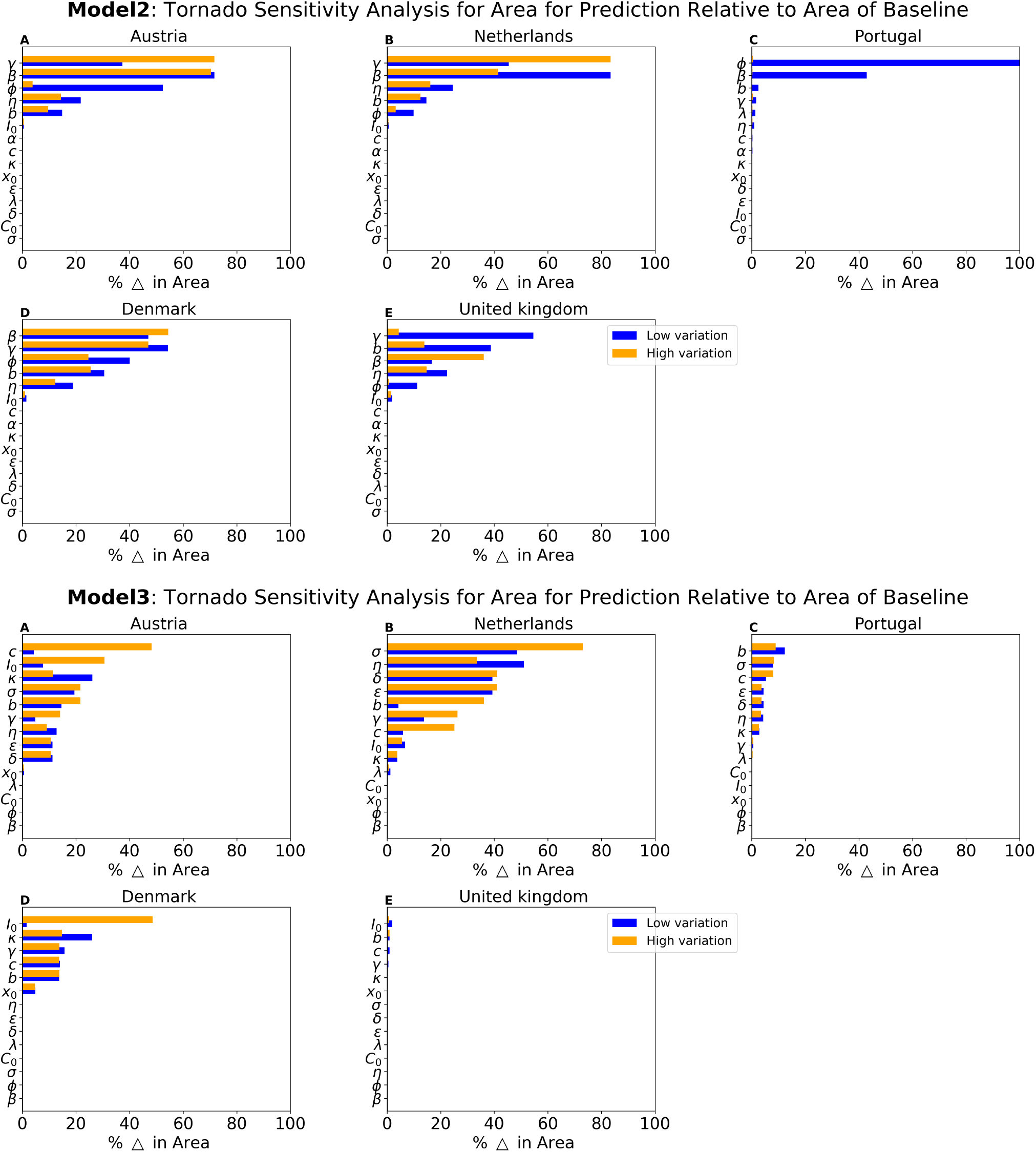
OAT sensitivity analysis using the percentage change in area for Model 2 and 3: One-at-a-time sensitivity analysis of each parameter in Model 2 and 3 where the area between the model’s incidence prediction and case notifications is calculate. The percentage change is calculated for each variation.

**Figure 11.**
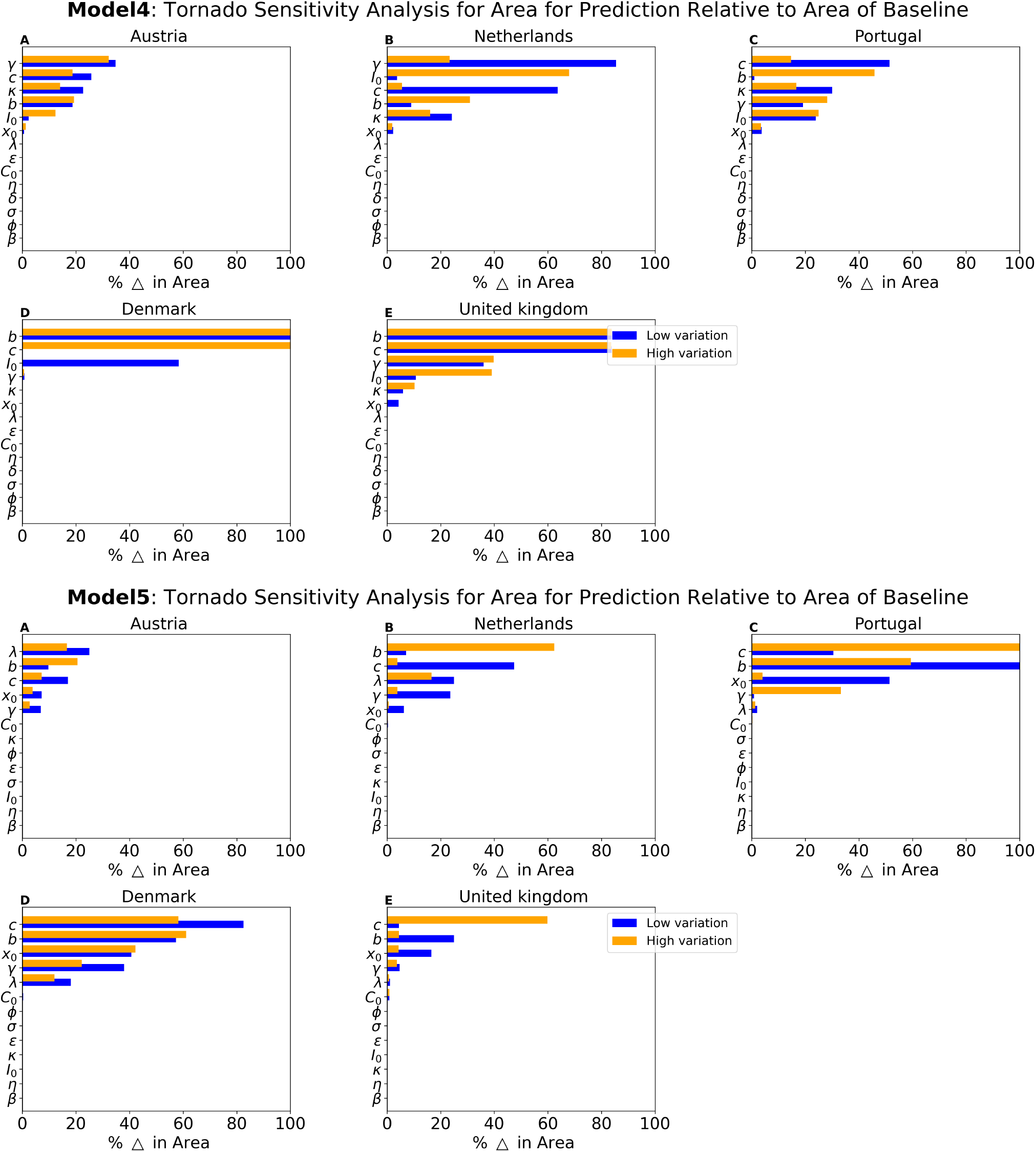
OAT sensitivity analysis using the percentage change in area for Model 4 and 5: One-at-a-time sensitivity analysis of each parameter in Model 4 and 5 where the area between the model’s incidence prediction and case notifications is calculate. The percentage change is calculated for each variation.

**Figure 12.**
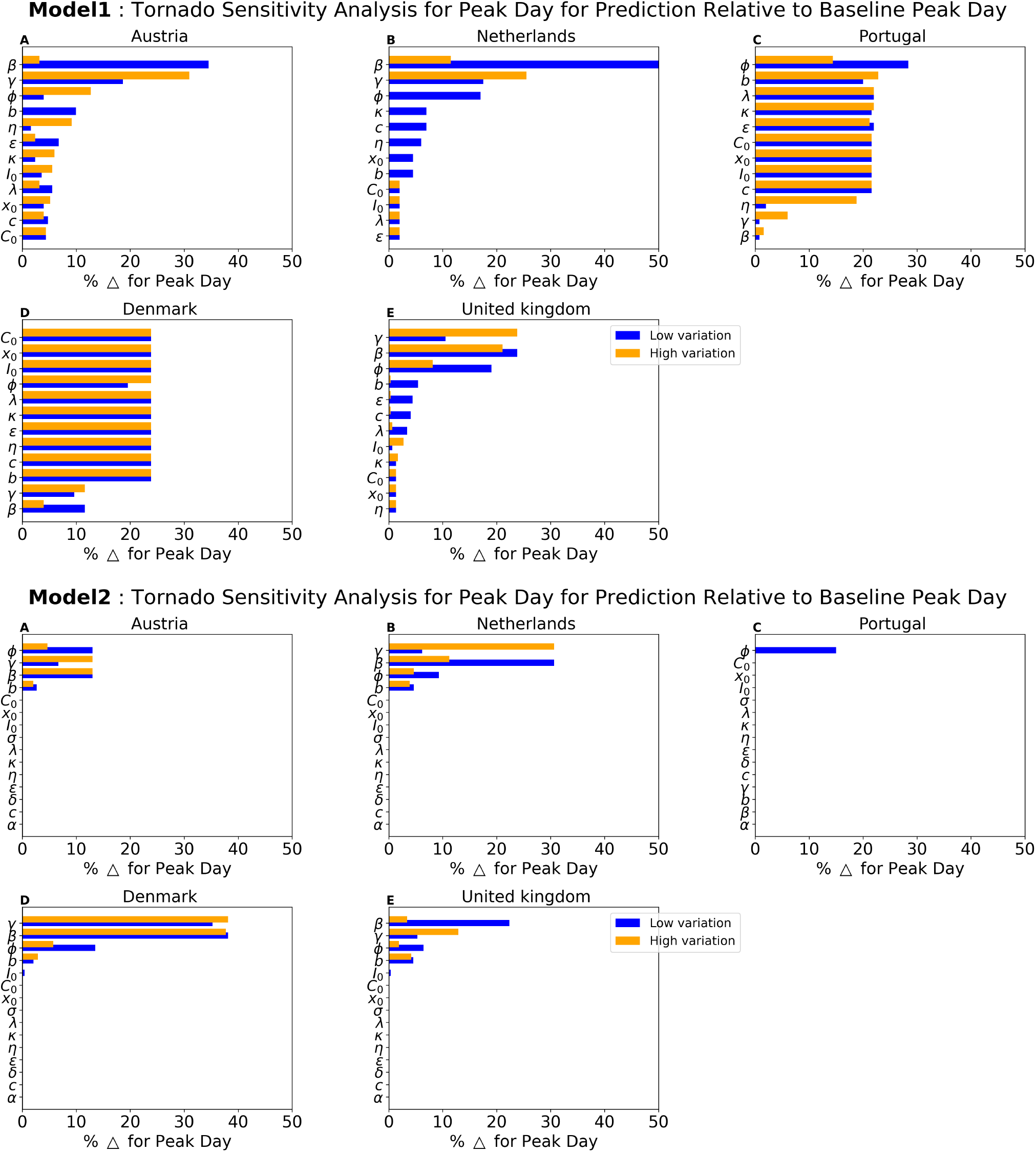
OAT sensitivity analysis using the percentage change in predicted peak day for Model 1 and 2: One-at-a-time sensitivity analysis of each parameter in Models 1 and 2 where we identify the peak day of the second wave. The percentage change of the peak days relative to the baseline is calculated for each variation.

**Figure 13.**
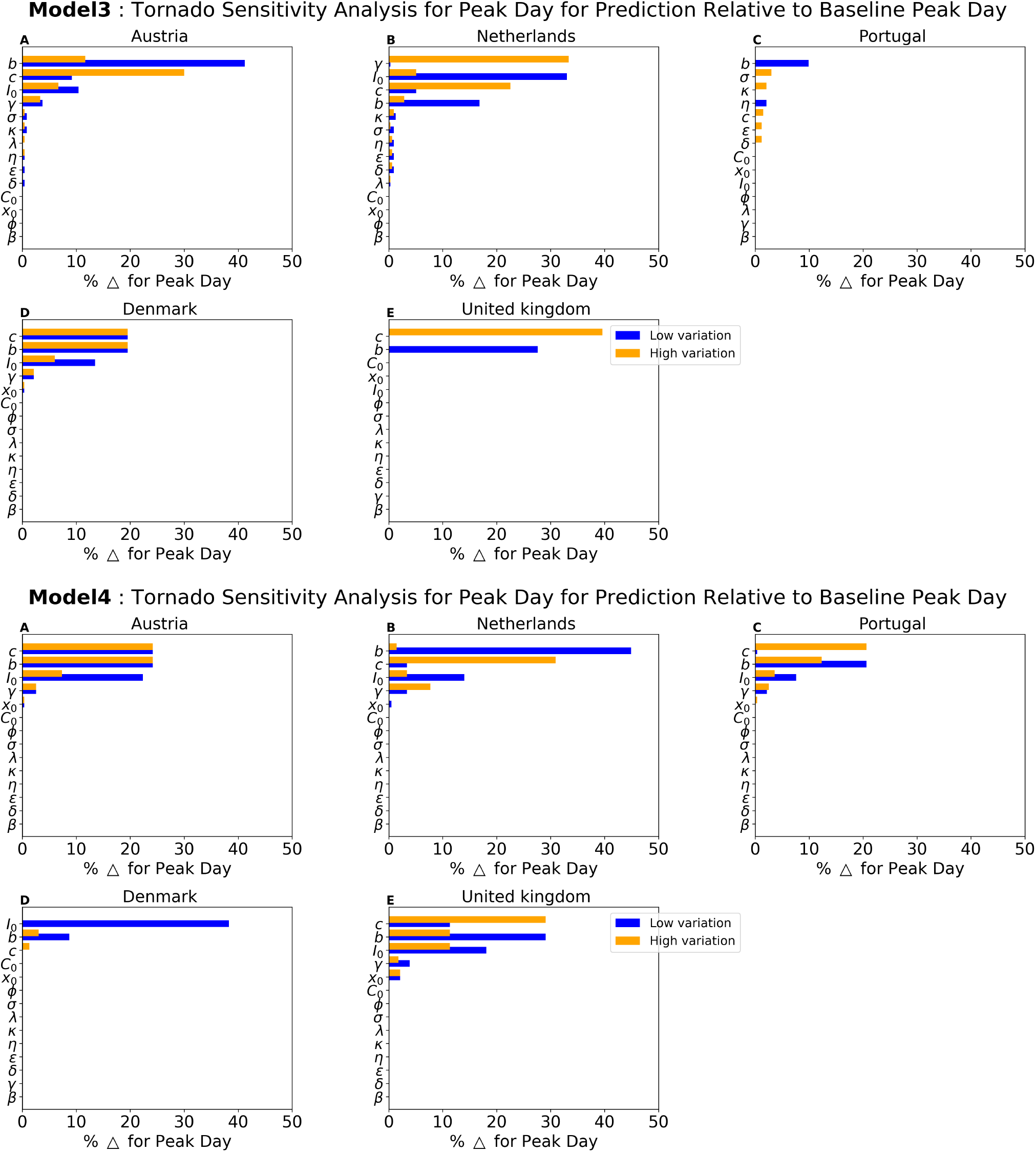
OAT sensitivity analysis using the percentage change in predicted peak day for Model 3 and 4: One-at-a-time sensitivity analysis of each parameter in Models 3 and 4 where we identify the peak day of the second wave. The percentage change of the peak days relative to the baseline is calculated for each variation.

**Figure 14.**
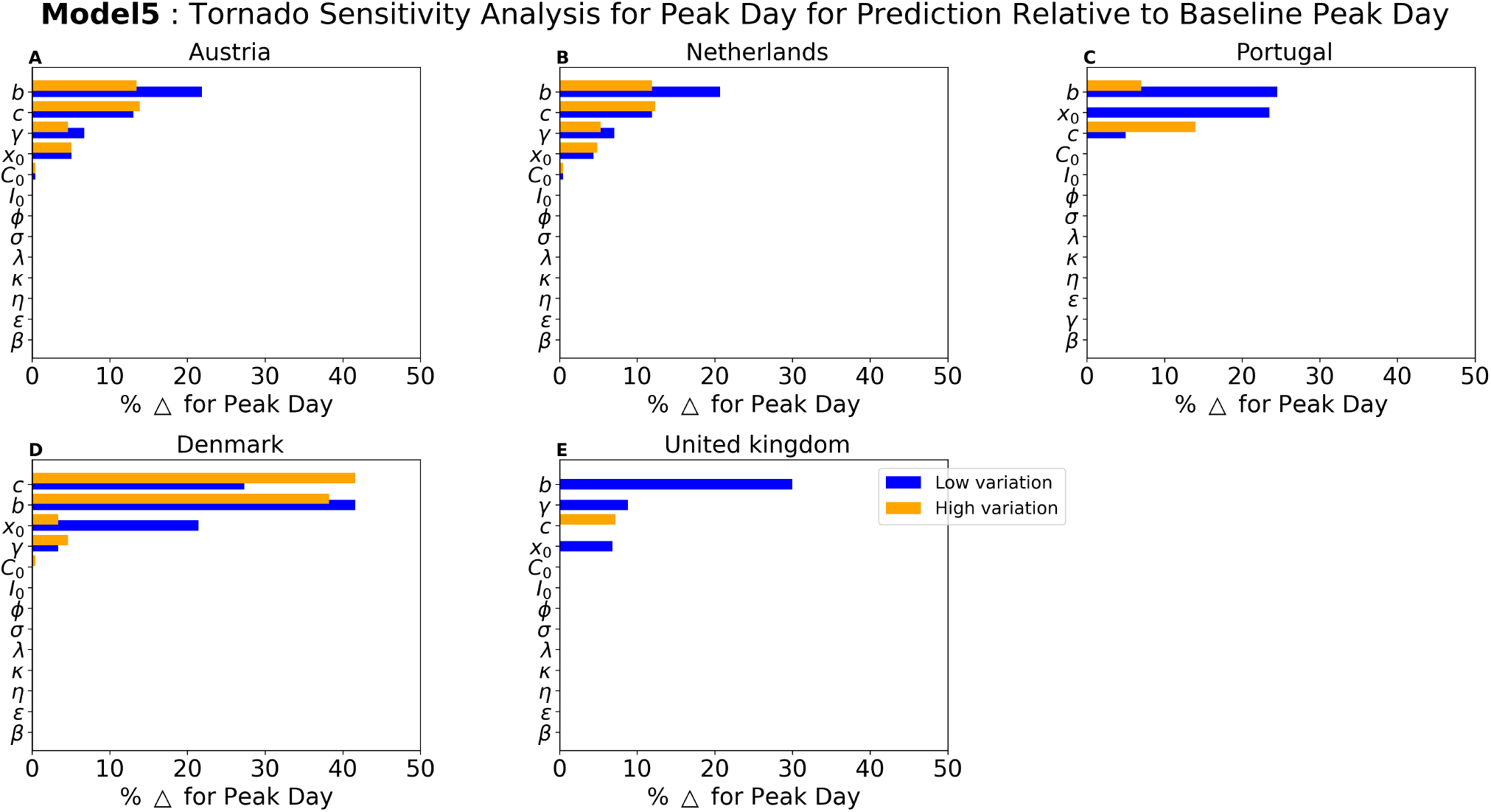
OAT sensitivity analysis using the percentage change in predicted peak day for Model 5: One-at-a-time sensitivity analysis of each parameter in Model 5 where we identify the peak day of the second wave. The percentage change of the peak days relative to the baseline is calculated for each variation.

**Figure 15.**
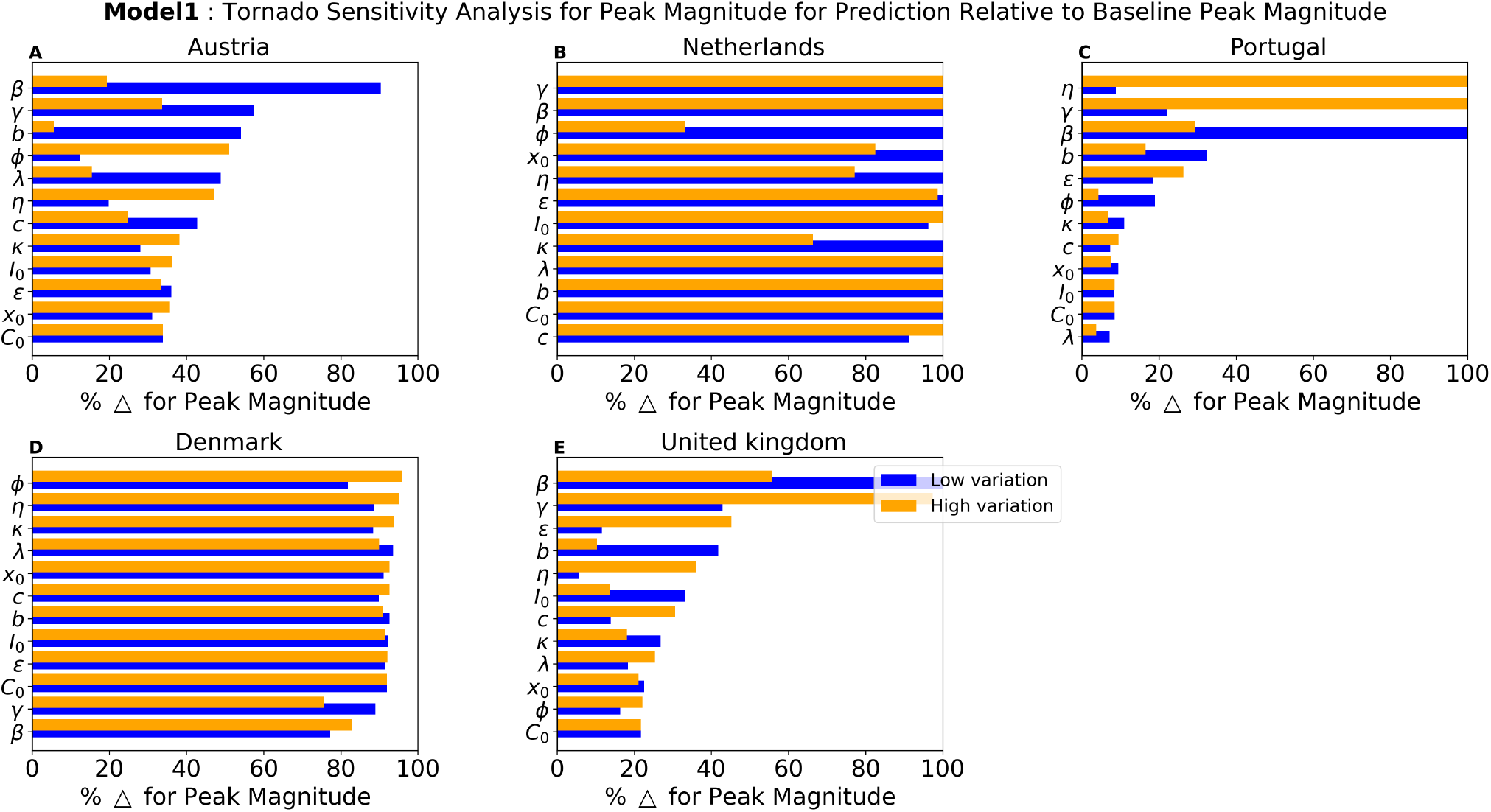
OAT sensitivity analysis using the percentage change in predicted peak magnitude for Model 1: One-at-a-time sensitivity analysis of each parameter in Model 1 where we identify the peak magnitude of the second wave. The percentage change of the peak magnitude relative to the baseline is calculated for each variation.

**Figure 16.**
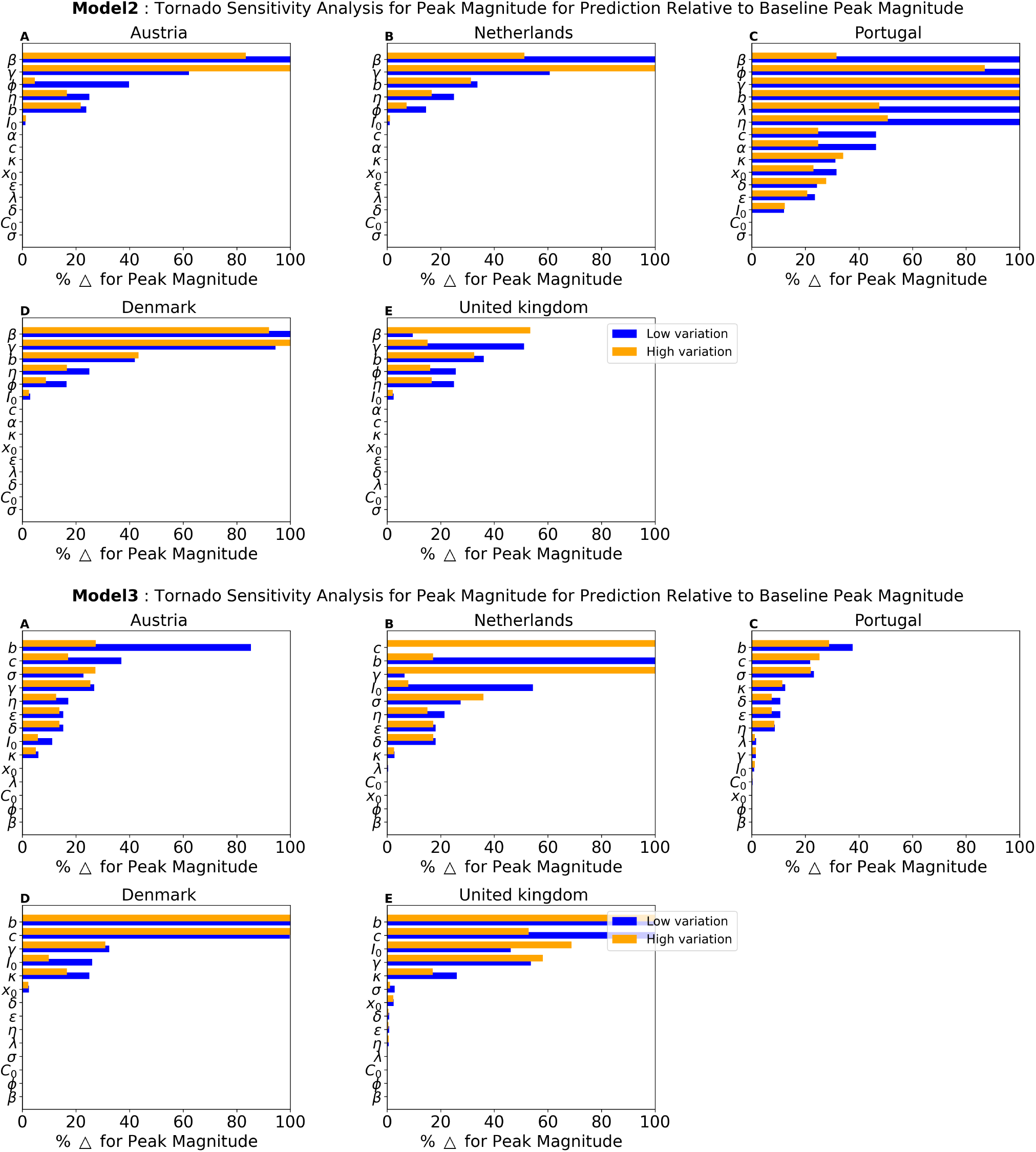
OAT sensitivity analysis using the percentage change in predicted peak magnitude for Models 2 and 3: One-at-a-time sensitivity analysis of each parameter in Models 2 and 3 where we identify the peak magnitude of the second wave. The percentage change of the peak magnitude relative to the baseline is calculated for each variation.

**Figure 17.**
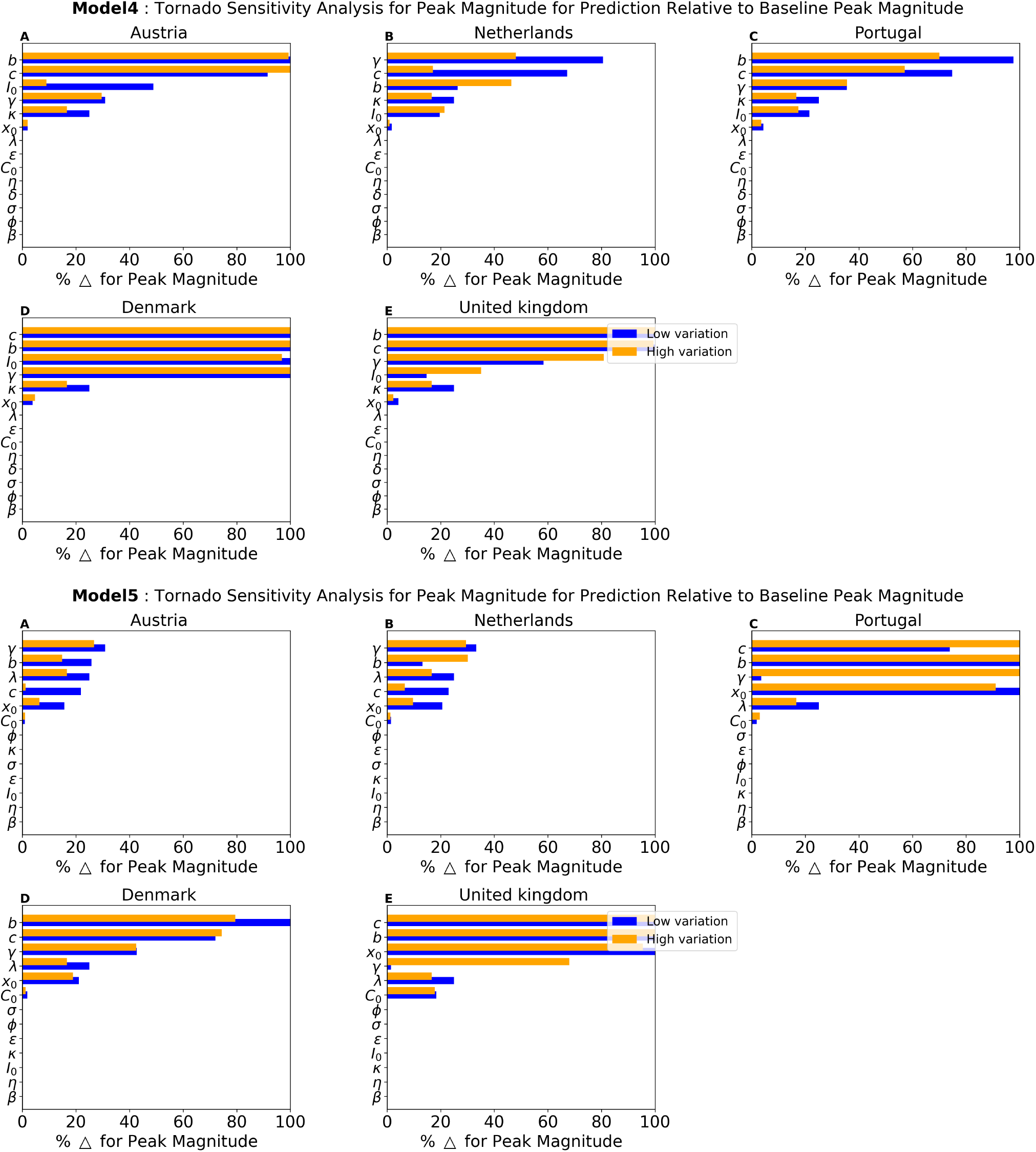
OAT sensitivity analysis using the percentage change in predicted peak magnitude for Models 4 and 5: One-at-a-time sensitivity analysis of each parameter in Models 4 and 5 where we identify the peak magnitude of the second wave. The percentage change of the peak magnitude relative to the baseline is calculated for each variation.

**Figure 18.**
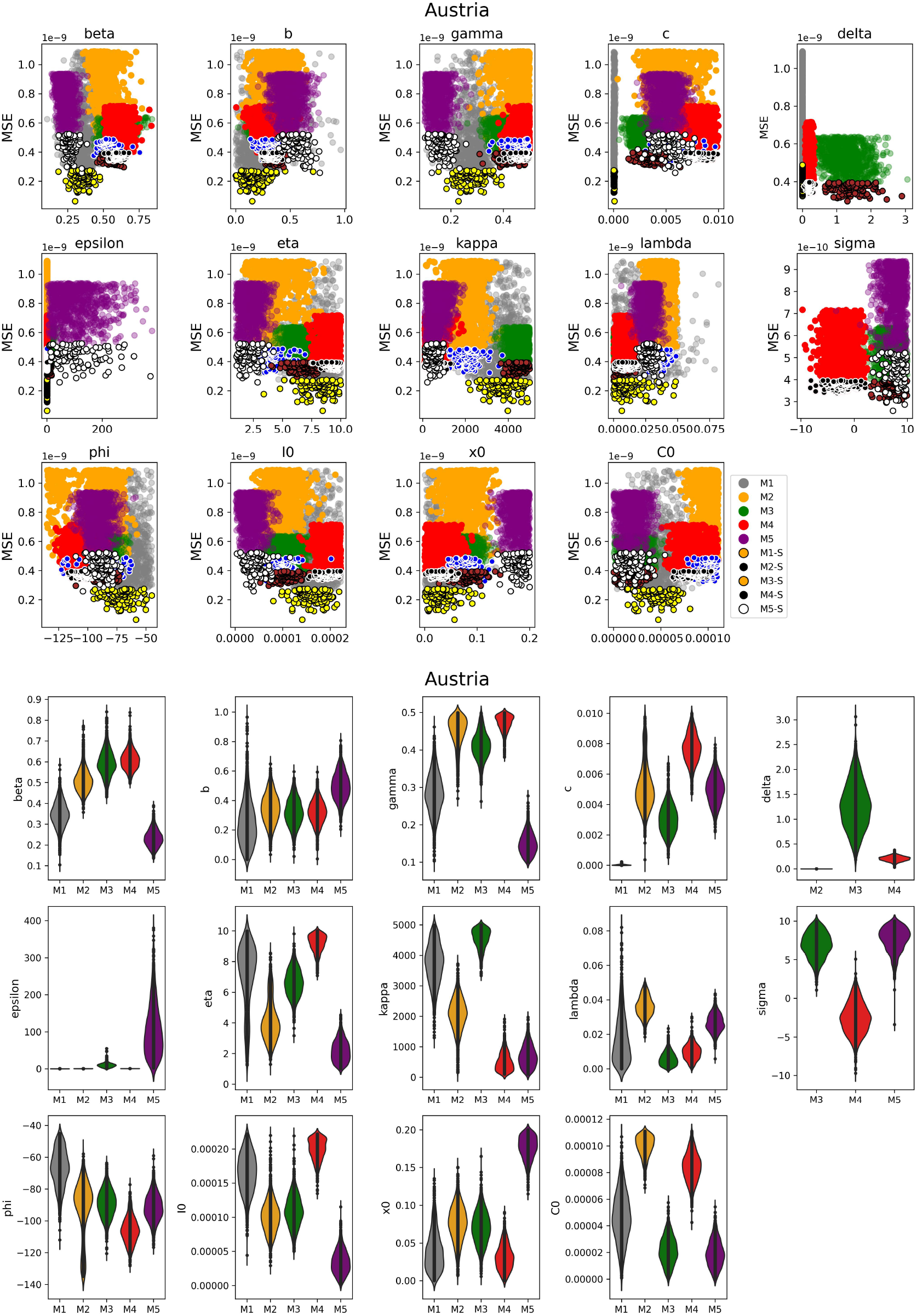
Posterior distributions on the inferred parameters for both models. Distribution of parameters against the mean squared error of 1000 particles and selected 100 particles used for the analysis. And a violin plot (right figure) which shows the type of distribution of the particles.

**Figure 19.**
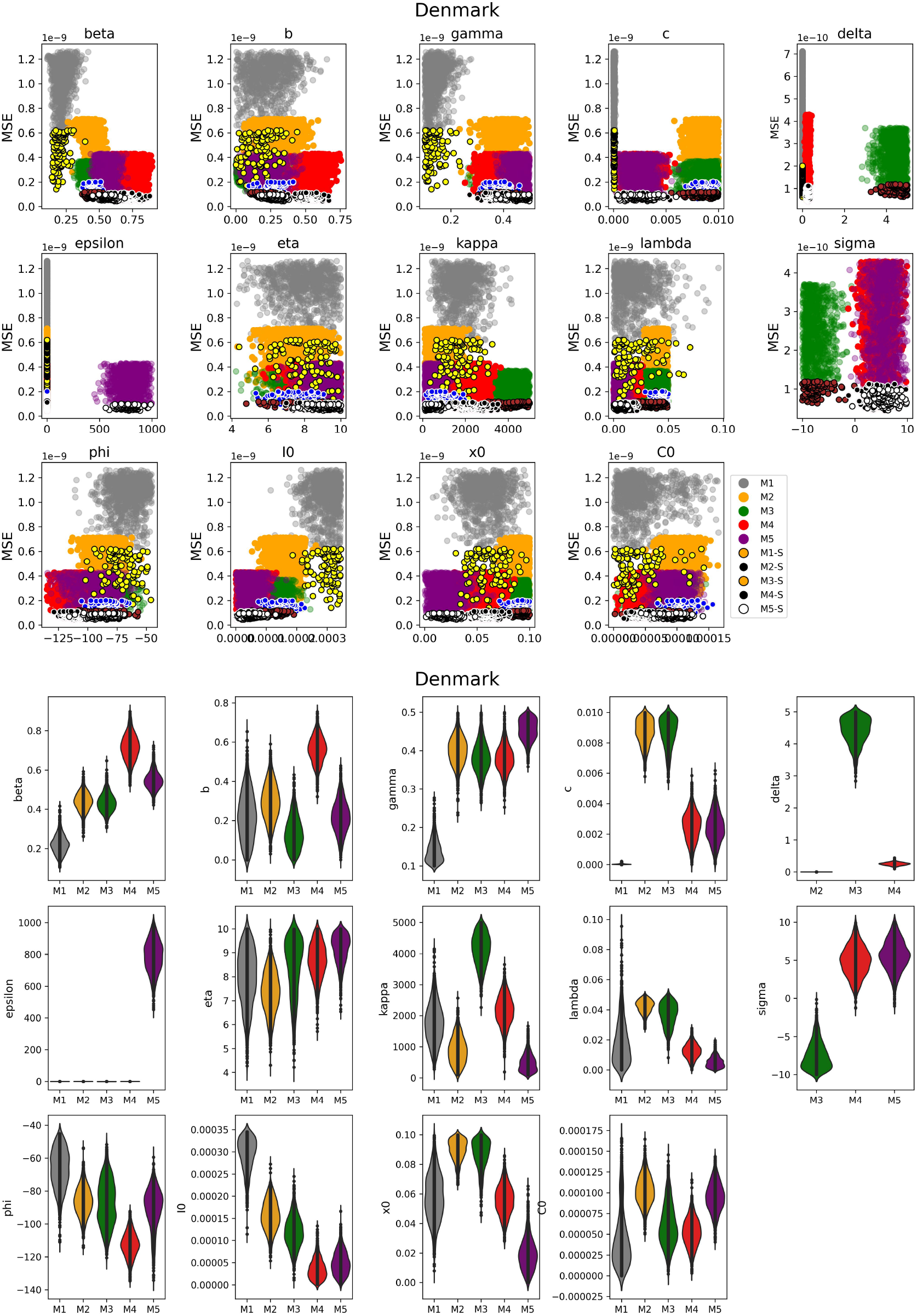
Posterior distributions on the inferred parameters for both models. Distribution of parameters against the mean squared error of 1000 particles and selected 100 particles used for the analysis. And a violin plot (right figure) which shows the type of distribution of the particles.

**Figure 20.**
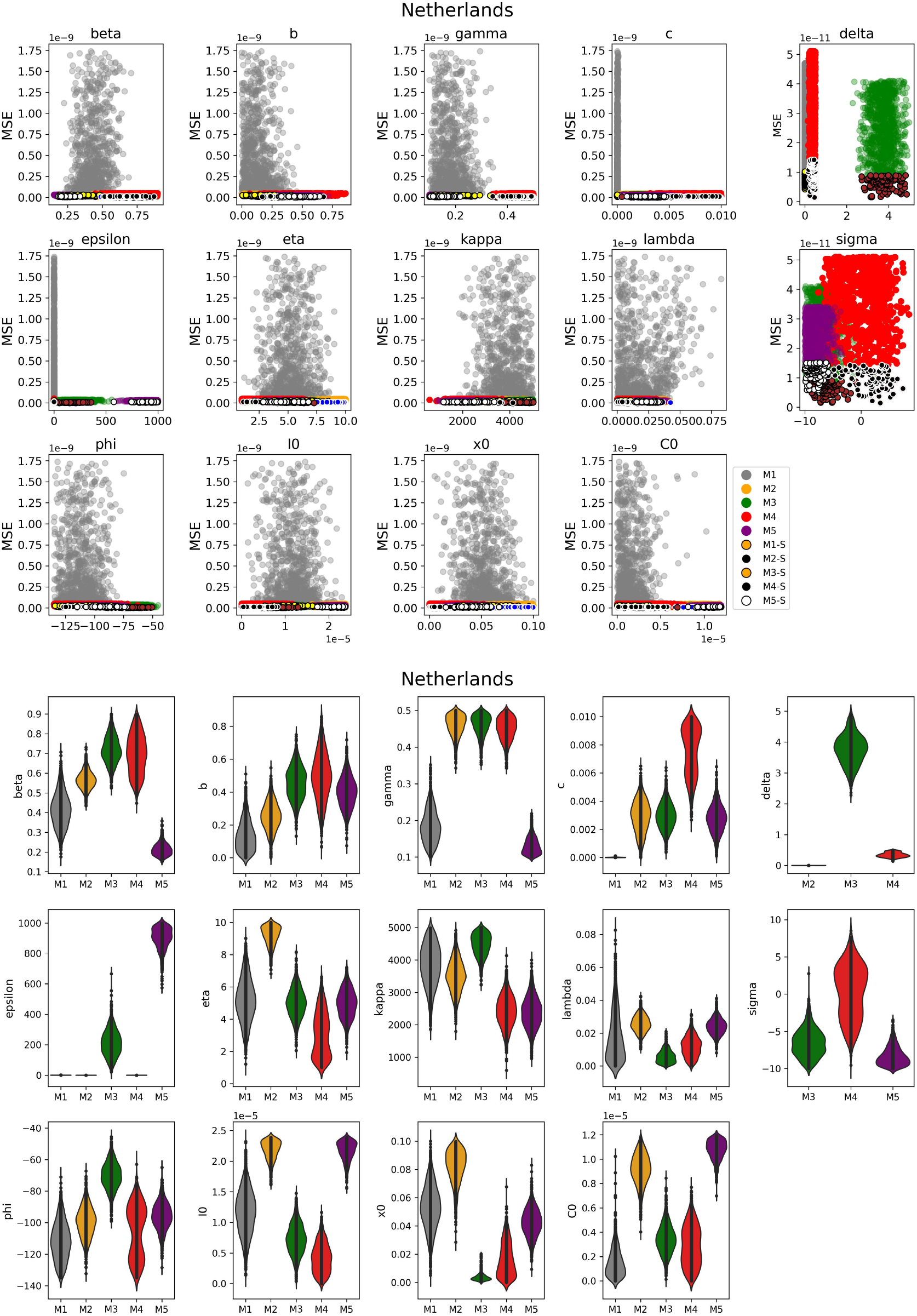
Posterior distributions on the inferred parameters for both models. Distribution of parameters against the mean squared error of 1000 particles and selected 100 particles used for the analysis. And a violin plot (right figure) which shows the type of distribution of the particles.

**Figure 21.**
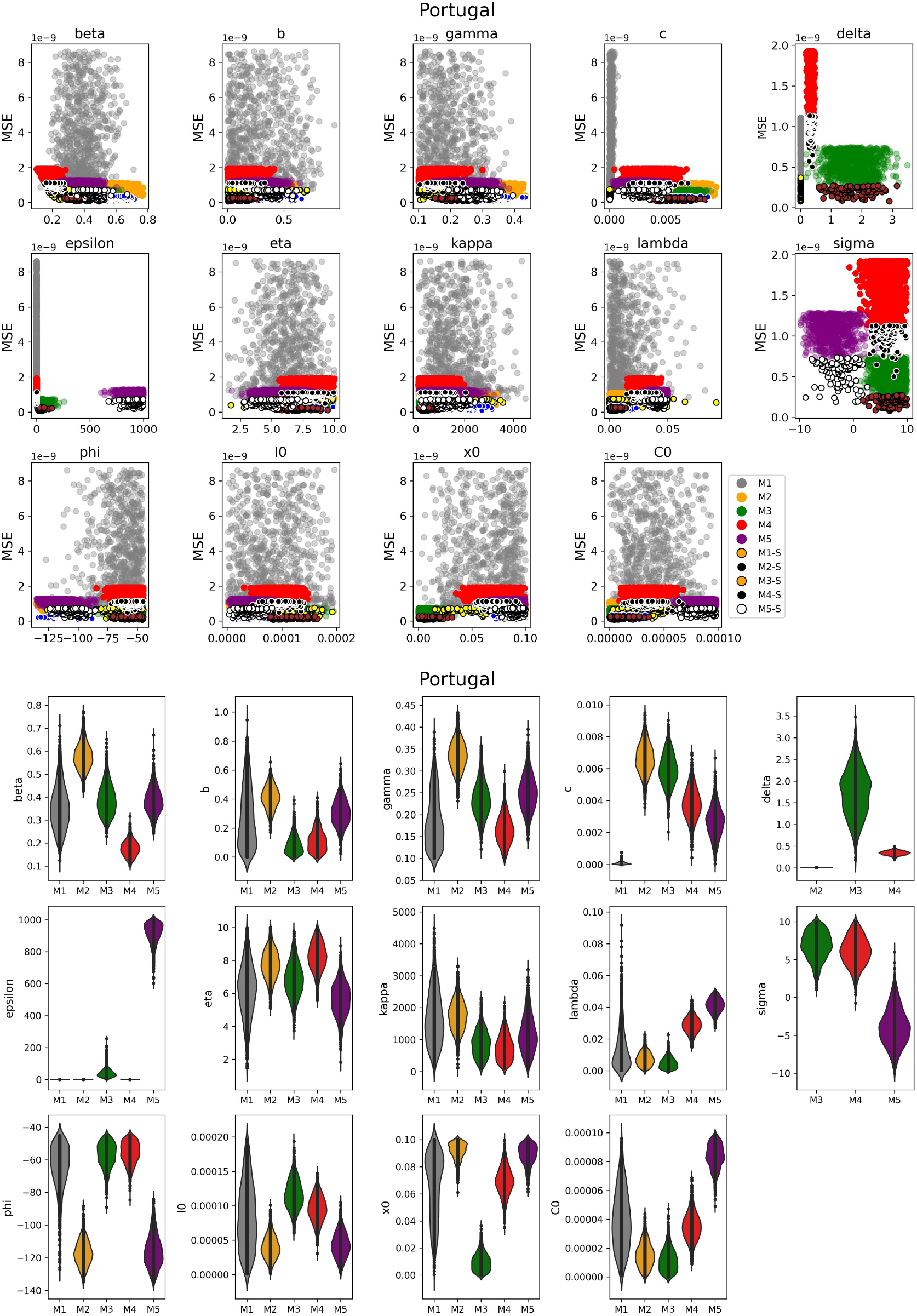
Posterior distributions on the inferred parameters for both models. Distribution of parameters against the mean squared error of 1000 particles and selected 100 particles used for the analysis. And a violin plot (right figure) which shows the type of distribution of the particles.

**Figure 22.**
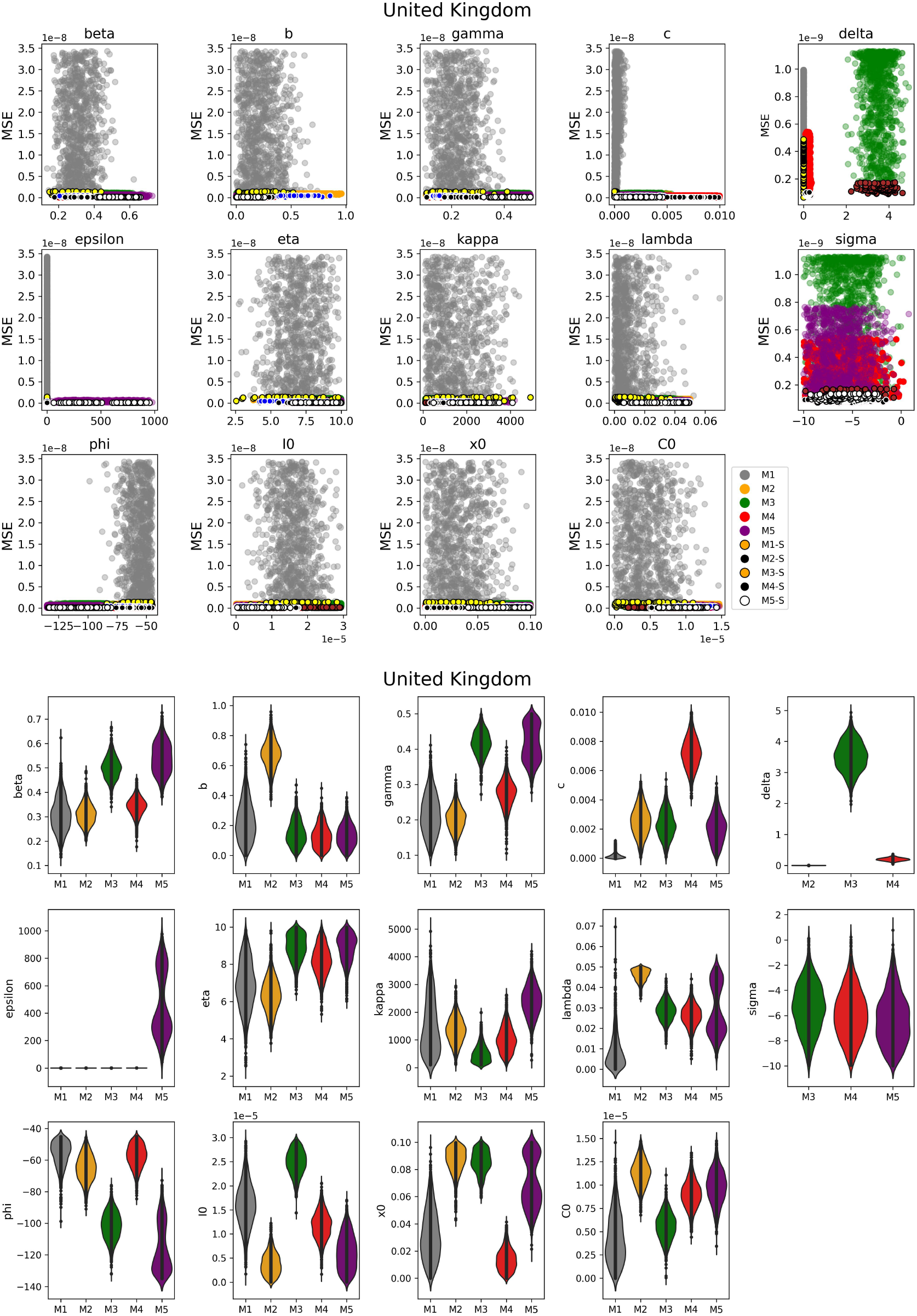
Posterior distributions on the inferred parameters for both models. Distribution of parameters against the mean squared error of 1000 particles and selected 100 particles used for the analysis. And a violin plot (right figure) which shows the type of distribution of the particles.

Interestingly, using Model 1 as a base model reference, which has 12 parameters, we observed that the two extra parameters (*α, δ*_2_) included in Model 2 had no influence on the area, peak day, or peak magnitude. Model 3, on the other hand, demonstrated a high level of sensitivity to the extra parameter (*δ*_3_). Model 4 does not record any sensitivity for the additional parameter (*δ*_4_). Since the structure of Model 5 differs from the others, we will not attempt to compare; however, we observed that *ε*_5_ *>* 1 was not sensitive to the model, but the initial proportion of mitigators was quite sensitive.

### 1.2 Particle Analysis

We estimate about 30,000 particles (parameter sets) for 1,000 particles per iteration (population) for each country and model. We selected 100 particles out of the 1,000 particles for our analysis (see main text **??**). The performance of the fitting algorithm is shown here with respect to their convergence and the identified iteration with the lowest error measure (mean squared error).

### 1.3 Parameter Identifiability

We observe from the posterior that it either has a wide distribution for most parameters or a narrow, peaked distribution. These could imply weakly identifiable parameters for the former, and as such, the empirical might be a limitation of the needed information. We identified that the rank for the Fisher Information Matrix (FIM) was 0 *< r* (number of model parameters) for Models 2 to 5 for all five countries using one identified good fit parameter value. However, for Model 1, we obtained the following results for the rank: Austria - 8, the Netherlands - 8, Portugal - 9, Denmark - 8, and the United Kingdom - 8. The correlation matrix shows a strong correlation between parameter pairs for Model 2 for all countries, Model 3 for the Netherlands, and Model 5 for the United Kingdom, unlike Model 1.

